# Multi-ancestry genome-wide association meta-analysis of mosaic loss of chromosome Y in the Million Veteran Program identifies 240 novel loci

**DOI:** 10.1101/2024.04.24.24306301

**Authors:** Michael Francis, Bryan R. Gorman, Tim B. Bigdeli, Giulio Genovese, Georgios Voloudakis, Jaroslav Bendl, Biao Zeng, Sanan Venkatesh, Chris Chatzinakos, Gabriel Hoffman, Erin McAuley, Sun-Gou Ji, Kyriacos Markianos, Patrick A. Schreiner, Elizabeth Partan, Yunling Shi, Poornima Devineni, VA Million Veteran Program, Jennifer Moser, Sumitra Muralidhar, Rachel Ramoni, Alexander G. Bick, Pradeep Natarajan, Themistocles L. Assimes, Philip S. Tsao, Derek Klarin, Catherine Tcheandjieu, Neal S. Peachey, Sudha K. Iyengar, Panos Roussos, Saiju Pyarajan

**Affiliations:** Center for Data and Computational Sciences (C-DACS), VA Cooperative Studies Program, VA Boston Healthcare System, Boston, MA, 02130, USA; Booz Allen Hamilton, McLean, VA, 22102, USA; Research Service, VA New York Harbor Healthcare System, Brooklyn, NY, 11209, USA; Department of Psychiatry and Behavioral Sciences, SUNY Downstate Health Sciences University, Brooklyn, NY, 11203, USA; Program in Medical and Population Genetics, Broad Institute of MIT and Harvard, Cambridge, MA, 02142, USA; Stanley Center, Broad Institute of MIT and Harvard, Cambridge, MA, 02142, USA; Department of Genetics, Harvard Medical School, Boston, MA, 02115, USA; Center for Precision Medicine and Translational Therapeutics; VISN 2 Mental Illness Research, Education, and Clinical Center (MIRECC), James J Peters Veteran Affairs Medical Center, New York/New Jersey VA Health Care Network, Bronx, NY, 10468, USA; Center for Disease Neurogenomics; Department of Psychiatry; Friedman Brain Institute; Department of Genetics and Genomic Sciences, Icahn School of Medicine at Mount Sinai, New York, NY, 10029, USA; Center for Data and Computational Sciences (C-DACS), VA Cooperative Studies Program, VA Boston Healthcare System, Boston, MA, USA; Center for Data and Computational Sciences (C-DACS), VA Boston Healthcare System, Boston, MA, USA; Office of Research and Development, Department of Veterans Affairs, Washington, DC, 20420, USA; Division of Genetic Medicine, Vanderbilt University Medical Center, Nashville, TN, 37232, USA; Department of Medicine, Cardiology Division, Massachusetts General Hospital, Boston, MA, 02114, USA; Cardiovascular Research Center, Massachusetts General Hospital, Boston, MA, 02114, USA; VA Palo Alto Epidemiology Research and Information Center for Genomics, VA Palo Alto Health Care System, Palo Alto, CA, 94304, USA; Department of Medicine, Stanford University School of Medicine, Stanford, CA, 94305, USA; Stanford Cardiovascular Institute, Stanford University School of Medicine, Stanford, CA, 94305, USA; VA Palo Alto Health Care System, Palo Alto, CA, USA; Department of Surgery, Stanford University School of Medicine, Stanford, CA, USA; VA Palo Alto Health Care System; Department of Medicine, Division of Cardiovascular Medicine, Stanford University School of Medicine, Stanford, CA, USA; Gladstone Institute of Data Science and Biotechnology, Gladstone Institutes, San Francisco, CA, USA; Department of Epidemiology and Biostatistics, University of California San Francisco, San Francisco, CA, USA; Research Service, VA Northeast Ohio Healthcare System, Cleveland, OH, 44106, USA; Cole Eye Institute, Cleveland Clinic, Cleveland, OH, 44195, USA; Department of Ophthalmology, Cleveland Clinic Lerner College of Medicine of Case Western Reserve University, Cleveland, OH, 44195, USA; Department of Population and Quantitative Health Sciences, Case Western Reserve University, Cleveland, OH, 44106, USA; Department of Ophthalmology and Visual Sciences, University Hospitals Eye Institute, Cleveland, OH, 44106, USA; Department of Genetics & Genome Sciences, Case Western Reserve University, Cleveland, OH, 44106, USA; Cleveland Institute for Computational Biology, Case Western Reserve University, Cleveland, OH, 44106, USA; Department of Medicine, Brigham and Women’s Hospital, Harvard Medical School, Boston, MA, 02115, USA

## Abstract

Mosaic loss of chromosome Y (mLOY) is the most commonly detected mosaic chromosomal alteration, and is associated with a range of health outcomes in males. We detected mLOY in 106,054 European (EUR), 13,927 admixed African (AFR), and 6,127 Hispanic (HIS) participants of the Million Veteran Program (MVP; N_total_=544,112). mLOY was positively associated with cigarette smoking, and negatively with obesity and type 2 diabetes. Multi-ancestry genome-wide association meta-analysis of MVP and UK Biobank mLOY summary statistics, jointly comprising 167,899 cases and 581,224 controls, identified 380 independent risk loci–240 of which were novel. GWAS in MVP high cell fraction (CF>10%) mLOY cases exhibited stronger effect sizes across risk loci. Local-ancestry-aware GWAS resolved ancestry-specific signals at *BCL2L1* and *SETBP1*. Integrative eQTL analyses highlighted 51 genes that causally influence mLOY via differential expression. Finally, traits with genetic correlations and polygenic scores significantly associated with mLOY were selected for Mendelian randomization (MR), implicating thirteen traits as causal influences on mLOY, including triglycerides, high-density lipoprotein, and body mass index.

## Introduction

Mosaic loss of chromosome Y (mLOY, also abbreviated LOY) is the most commonly detected mosaic chromosomal alteration (mCA)^1–4^. mLOY is readily detected in leukocytes^5^, which are the primary source of blood-derived DNA, and is observable in upwards of 40% of males above age 70^1,6^, and 70% of males over 85^7^. High turnover in the hematopoietic stem cell compartment drives clonal expansion in mosaic cell subpopulations possessing selective advantages, though it is unknown if mLOY itself contributes to this selection^5,8,9^.

Historically, the gene-poor and repetitive-element-rich composition of the Y chromosome led researchers to believe its role was restricted to spermatogenesis and sex determination^10^. Congruently, mLOY was initially considered to be a benign condition, and a consequence of the broader genomic instability that occurs with aging^11^. In the past decade, epidemiological studies have reported associations between mLOY and a wide range of health outcomes^5^, including positive associations with all-cause mortality^12–14^, hematological malignancies and other cancers^12,13,15^, Alzheimer’s disease^16^, and cardiovascular disease (CVD)^5,12,17^; negative associations with body mass index (BMI)^17^ and gout^17^; and mixed associations with type 2 diabetes (T2D)^12,17^. These observed associations with comorbidities have also been inconsistent across studies^5^, perhaps owing to differences in sample collection and mLOY classification methods, such as low versus high cell fraction (CF) detection^6^. Importantly, it has yet to be determined whether mLOY is a driving causal factor of these conditions, a “passenger” (i.e., a consequence), or is symptomatic of a shared, underlying cause, such as genetic susceptibility to DNA replication errors, or exogenous exposure to mutagens (particularly via smoking cigarettes)^5,18^.

The mechanisms by which mLOY contributes to pathogenesis are potentially wide-ranging and gradually being uncovered. For example, in a murine model of mLOY, fibrotic deposition in the extracellular matrix was shown to induce CVD^14^. In another study, CRISPR–Cas9 deletion of chromosome Y produced more aggressive bladder cancer by means of T-cell exhaustion^19^. It has also been suggested that cell lineage influences the relationship between mLOY and transcriptional dysregulation, which in turn produces distinct disease phenotypes^18^.

Estimates of single nucleotide polymorphism-based heritability (SNP-*h*^2^) for mLOY detected via the pseudo-autosomal region 1 (PAR1) are as high as 31.7%, highlighting mLOY as substantially heritable compared to most human traits^1^. Germline genetics govern many processes which can lead to somatic mCA acquisition and clonal expansion, particularly via genes involved in cell-cycle regulation, DNA damage response, apoptosis, and cancer susceptibility^1,8^. Advances in leveraging genotype phase to detect subtle imbalances in allelic ratio have enabled sensitive and accurate identification of mLOY in large-scale cohorts^1^. A genome-wide association study (GWAS) of mLOY status, performed in European ancestry (EUR) UK Biobank (UKB) participants^1^, replicated all 19 previously reported genetic risk loci^20^, including the oncogene *TCL1A*^15^, and identified 137 novel loci. A GWAS of mLOY in BioBank Japan (BBJ), which used the mean logarithm of R ratio probe intensity in chromosome Y (mLRR-Y) as a quantitative proxy for mean Y chromosome dosage, identified 50 signals in 46 loci, 35 of which were reported as novel^21^. Subsequent mLOY GWAS did not reveal additional novel loci^22,23^.

In this study we analyzed 544,112 male participants (126,108 cases and 418,004 controls) from the Million Veteran Program (MVP), a biobank in the Department of Veterans Affairs (VA) healthcare system which combines ancestrally diverse genetic data with extensive electronic health records (EHRs)^24^. In addition to a large EUR cohort, we present the first GWAS of mLOY status in African (AFR) and Hispanic/Latino (HIS) ancestries. We then meta-analyzed the MVP GWAS results with a previous GWAS from UKB^1^, producing the largest mLOY study to-date (N=749,123). MVP provides a uniquely valuable resource to perform multi-ancestry mLOY analyses, as it avoids technical issues related to genotyping and mosaicism calling that may be introduced by combining data from separate biobanks. Additionally, our results highlight the benefits of inclusive population studies in advancing our understanding of mLOY.

## Results

### mLOY phenotyping and prevalence across ancestries

In all analyses performed in this study (Fig. 1) we classified mLOY cases in MVP as having any detectable CF of mosaicism in chromosome Y. MoChA^8,25^ was used to detect shifts in allelic ratios of phased PAR1 and PAR2 haplotypes shared by the X and Y chromosomes (Fig. 2a), similar to a previous mLOY GWAS in UKB^1^. In MVP EUR, 106,054 of 400,970 men (26.4%; median age 66) had detectable mLOY. Lower prevalences were observed in MVP AFR (13,927 of 99,103; 14.1%; median age 60) and MVP HIS (6,127 of 44,039; 13.9%; median age 60) (Supplementary Data 1). As expected, mLOY prevalence increased with age, from 10% among participants aged 50-60 to upwards of 50% in octogenarians (Fig. 2b-c). CF also increased with age across all ancestries (Extended Data Fig. 1a); high CF mLOY (>10%) was observed in 56%, 37%, and 44% of cases in EUR, AFR, and HIS, respectively (Extended Data Fig. 1b).

**Fig. 1.**
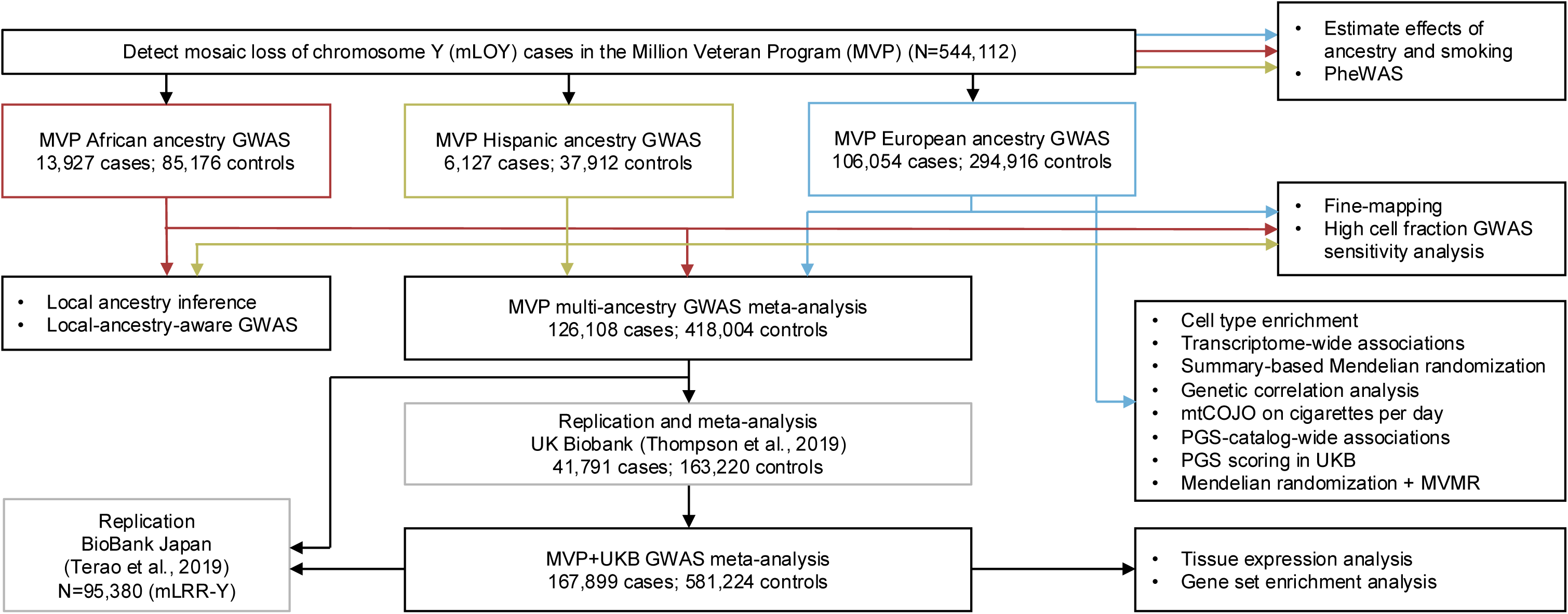
Overview of genome-wide association study for mosaic loss of chromosome Y (mLOY) in Million Veteran Program (MVP) participants. Genome-wide association studies (GWAS) were conducted in MVP European, African, and Hispanic ancestries. These were then meta-analyzed with UK Biobank^1^. Replication was conducted using UK Biobank^1^ and BioBank Japan^21^. Several follow-up functional analyses were performed, indicated by bulleted lists. BMI, body mass index; eQTL, expression quantitative trait loci; GSEA, gene set enrichment analysis; mLRR-Y, mean logarithm of R ratio in chromosome Y; mtCOJO, multi-trait-based conditional & joint association analysis; MVMR, multivariable Mendelian randomization; PGS, polygenic score; PheWAS, phenome-wide association study.

**Fig. 2.**
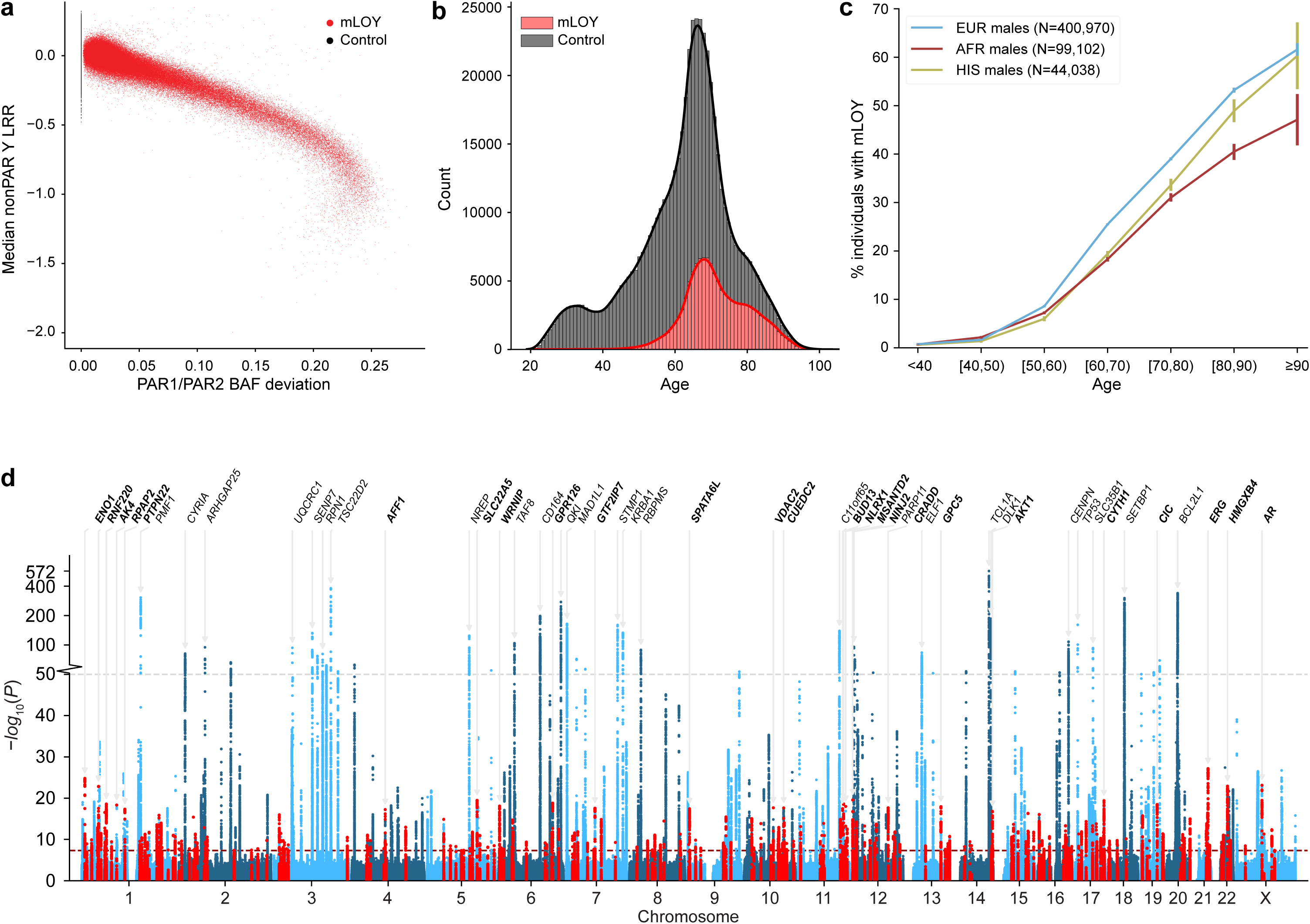
Mosaic loss of chromosome Y (mLOY) in the Million Veteran Program (MVP) **a**, Median genotyping probe intensity log R ratio (LRR) vs. phased B Allele Frequency (BAF) in the pseudo-autosomal regions (PAR) 1 and 2. **b**, Density of age distribution in all MVP mLOY cases and controls. **c**, Percentage of individuals with mLOY per ten-year age bin for MVP European (EUR), African (AFR), and Hispanic (HIS) cohorts. Error bars represent 95% confidence intervals. **d**, Manhattan plots show the −log_10_(*P*) for associations of genetic variants with mLOY in the multi-ancestry meta-analysis. Novel mLOY index variants and surrounding variants within ±50 Kb are highlighted in red. The red line indicates the genome-wide significance threshold (*P*<5×10^−8^). The grey dotted line represents a transition from linear to log-scale on the y-axis. The 25 most significant novel (bold) and known loci are annotated with the nearest gene.

To evaluate the impact of ancestry on mLOY prevalence within the three MVP ancestry groups, which had varying age distributions, we generated sensitivity models with sequential covariate adjustment, and applied balancing weights to estimate effects using weighted samples (Extended Data Fig. 2a-c; Supplementary Data 2-3). Upon adjusting for age, age-squared, BMI, smoking status, interactions of these covariates with ancestry, and age*BMI interaction, we observed a reduced marginal odds ratio (OR) of mLOY in AFR (OR=0.72, 95% confidence interval = [0.71, 0.74]) and HIS (OR=0.81 [0.79, 0.84]) compared to EUR (Extended Data Fig. 2d; Supplementary Data 3). The consistency of these ORs in both weighted and unweighted models increase our confidence in the observed associations. Additionally these findings replicate the results of a recent study in TOPMed^23^. Using the same modeling strategy, we observed an increased association with mLOY in former smokers (OR=1.26 [1.24, 1.28]) and current smokers (OR=1.64 [1.59, 1.70]) (Extended Data Fig. 2e; Supplementary Data 3). Similarly, former and current smokers had higher mLOY CF in all ancestries (Extended Data Fig. 1c). These findings that smoking is associated with increased risk of mLOY are consistent with previous reports^12,15,21,23,26^.

### Phenome-wide associations between mLOY and comorbidity prevalence

We performed phenome-wide association studies (PheWAS) to identify associations between prevalent disease and mLOY (Supplementary Data 4-6). We observed 344 significant associations in EUR; top associations included increased tobacco use disorder (OR=1.65 [1.62, 1.68]) and chronic airway obstruction (OR=1.35 [1.32, 1.37]), and decreased obesity (OR=0.71 [0.70, 0.72]), T2D (OR=0.75 [0.74, 0.76], and gout (OR=0.68 (0.67, 0.70) (Extended Data Fig. 3a). Expectedly, the associations observed in the full EUR cohort related to respiratory traits and substance use were greatly attenuated in a stratified analysis of non-smokers; patterns of decreased obesity, diabetes, gout, and renal and heart diseases remained significant (Extended Data Fig. 3b). The strongest hematopoietic trait association was decreased thrombocytopenia (OR=0.70 [0.67, 0.73])^27^. Highly similar PheWAS results were observed in AFR and HIS (Supplementary Data 5-6; Supplementary Figs. 1-3), as well as in stratified analyses of current smokers and age-matched high CF cases (Supplementary Data 4-7).

### Genome-wide significant associations in MVP EUR, AFR, and HIS

We performed a GWAS for mLOY case-control status in each MVP ancestry group (Supplementary Data 8), and, using COJO (conditional and joint analysis), identified 336 conditionally independent association signals reaching genome-wide significance (GWS; *P*<5×10^−8^) in EUR, 50 in AFR, and 17 in HIS (Extended Data Fig. 4; Supplementary Fig. 4a-c; Supplementary Data 9-11). The most significant signals in EUR and HIS were in *TCL1A*; in EUR, we identified the same *TCL1A* lead risk allele as in previous GWAS^1,15^ (rs2887399-T; OR=0.71 [0.70, 0.72]; *P*=3.18×10^−419^; Supplementary Fig. 5a). The strongest effect observed in the MVP EUR GWAS, in variants with minor allele frequency (MAF) ≥0.1, was in *TP53* (rs78378222; OR=1.66 [1.58, 1.75]; P=1.44×10^−90^). Of the MVP EUR signals, 222 were within 1 Mb of a previously reported mLOY index variant^1,20,21^, including 148 of the 156 previously reported in UKB^1^, and effect sizes were highly concordant between these two European cohorts (*r*=0.91; Supplementary Fig. 6a). In AFR, the most significant association was in *RPN1* (rs113336380), which has a MAF of ∼6% in AFR and <0.01% in EUR (Supplementary Fig. 5b).

We improved our variant selection by fine-mapping and estimating credible sets of candidate causal variants in each ancestry. In EUR, we identified 11,242 variants in 334 high-quality credible sets, with a median of 8.5 variants per set (Supplementary Data 12); in AFR, 533 variants in 45 high-quality credible sets, with a median of 5 variants per set (Supplementary Data 13); and in HIS, 713 variants in 16 high-quality credible sets, with a median of 12.5 variants per set (Supplementary Data 14).

Effect directions were concordant between EUR and AFR associations (*r*=0.66, *P*=1.13×10^−46^), with the exception of a small group of novel variants that were exclusively found in EUR (Supplementary Fig. 6b). All GWS HIS signals were also GWS in EUR, with concordant effects (*r*=0.85; Supplementary Fig. 6c). In these three MVP single ancestry GWAS, we report 114 novel signals in EUR, and five additional GWS novel signals specific to AFR: rs2840287 (*MPL*), rs10786559 (*NKX2-3*), rs2724631 (*ETV6*), rs6018599 (*BLCAP*) and rs113887465 (*GGT5*/*CABIN1*) (Supplementary Fig. 7; Supplementary Data 9-10).

We extended our association analyses for all ancestries to a genotype panel enriched in protein-altering rare variants (MAF<0.1%)^28^, and identified four novel rare variants in EUR (Supplementary Data 15). These include a frameshift mutation at *DCXR*:c.583del (p.His195fs), and somatic missense mutations in *DNMT3A* (R882H)*, JAK2* (V617F), and *IDH2* (R140Q), which are known to be associated with hematologic malignancies such as clonal hematopoiesis of indeterminate potential (CHIP) and acute myeloid leukemia (AML)^29,30^. The estimated effects of these rare variants were negative and, consistent with expectation, stronger than those of common variants (Supplementary Fig. 8).

### High cell fraction GWAS

To explore whether higher CF cases of mLOY are driven by a unique genetic risk profile, we performed GWAS using only age-matched cases with CF>10% (Extended Data Fig. 4; Supplementary Fig. 4d-f; Supplementary Data 16-18). Interestingly, COJO signals in high CF GWAS had stronger effect sizes in a variant-level comparison with the full cohorts (Extended Data Fig. 4); this was consistent with expectations of a refined phenotypic signal in high CF cases. Overall, we observed strong correlations when comparing COJO signal effects across high CF and full cohorts (*r*=0.82 to 0.99; Supplementary Data 19). The high CF GWAS revealed two novel signals in AFR (Extended Data Fig. 4b; Supplementary Data 17). Additionally, there were three novel loci in HIS, however, we consider these to be lower confidence due because the lead variants were not replicated (*P*>0.05) in our other GWAS (Extended Data Fig. 4c; Supplementary Data 18).

### Meta-analyses of MVP and UKB

We performed a fixed-effects meta-analysis of MVP EUR, AFR, and HIS ancestries, for a total of 126,108 cases and 418,004 controls. This yielded 239 risk loci, 40 of which did not reach GWS in any individual MVP ancestry (Supplementary Fig. 9ab; Supplementary Data 20). We then performed a combined meta-analysis of MVP with UKB^1,31^, for a total of 749,123 samples (167,899 cases and 581,224 controls). In MVP+UKB, we identified 380 risk loci at GWS, 230 of which were novel in this study (Fig 2d; Supplementary Fig. 9c; Supplementary Data 21). Loci were defined using the two-stage clumping procedure implemented by the Functional Mapping and Annotation (FUMA) platform^32^. Locus novelty was defined as the absence of previously reported mLOY associations within the larger of either these locus boundaries or a ±1 Mb window. (The ±1 Mb window was also used to assess novelty of COJO index variants and rare variants). All novel loci in this study are represented in the MVP+UKB meta-analysis, with the exception of ten additional signals only found in a single GWAS (including the three rare variants), for a total of 240 novel mLOY loci (Supplementary Data 22).

Across MVP multi-ancestry and MVP+UKB meta-analysis index variants, rs2887399 (*TCL1A*) remained the most significant association (*P*_MVP+UKB_=7.93×10^−573^; Supplementary Fig. 5a). The strongest effects were observed at *CHEK2* (rs186430430; OR=1.78 [1.61, 1.97]) and *TP53* (rs78378222; OR=1.70 [1.64, 1.77]), whose gene products coordinate (together with *ATM*, which contains a rare variant associated with mLOY) in a crucial DNA damage response pathway. For both of these meta-analyses we also generated random-effects (RE) estimates using the Han-Eskin method (RE2)^33^, and observed similar *P*-values to FE (Supplementary Data 20-21).

### Replication in UK Biobank and BioBank Japan

We performed external replication of our GWAS results using the two previous largest mLOY GWAS, in UKB^1^ European and BBJ^21^ Japanese ancestry samples. We observed highly similar effects and *P*-values between MVP and UKB^1^. In a variant-level replication of the 327 MVP EUR signals that were available in UKB^1^, all but one (rs925301) had the same effect direction, and beta values were highly correlated (*r*=0.91; Supplementary Fig. 6a; Supplementary Data 9, 23). Of the 114 novel MVP EUR signals, 83 had *P*<0.01 and 16 had *P*<1×10^−5^ in UKB^1^. Highly similar replicability was observed between the predominantly European MVP multi-ancestry meta-analysis and UKB^1^ (Supplementary Data 20, 23).

We additionally queried BBJ^21^ for replicability using the 172 of 239 index variants from the MVP meta-analysis available in that dataset. Although BBJ used a less sensitive quantitative mLOY measure (mLRR-Y) as opposed to our case-control designation, the effects of 151 of 172 index variants were aligned, including all 71 variants with *P*<0.01 in BBJ (Supplementary Fig. 6d; Supplementary Data 20, 23). Finally, we compared our MVP+UKB meta-analysis with BBJ^21^, and found similar effect size and *P*-value concordance as with the MVP multi-ancestry meta-analysis (Supplementary Data 22-23). BBJ^21^ had 135 of 286 available MVP+UKB meta-analysis index variants (47.2%) with *P*<0.05 and shared 29 GWS signals (10%; Supplementary Data 22-23).

### Local-ancestry-aware associations in admixed groups

Next, we performed local ancestry inference^34^ in the two admixed MVP groups in this study, AFR and HIS. On average, AFR had 81.3% African and 18.7% European admixture; HIS had 58.1% European, 33.2% Native American (NAT), and 8.8% African admixture (Extended Data Fig. 5; Supplementary Data 25). We performed local-ancestry-aware GWAS using the Tractor model^35^ to resolve ancestry-specific mLOY association signals (Fig. 3a; Supplementary Figs. 10-11). Signals with large allele frequency differences between ancestries resulting from recent admixture can be separated with this method; we highlight two AFR loci at 20q11 and 18q12 to illustrate.

**Fig. 3.**
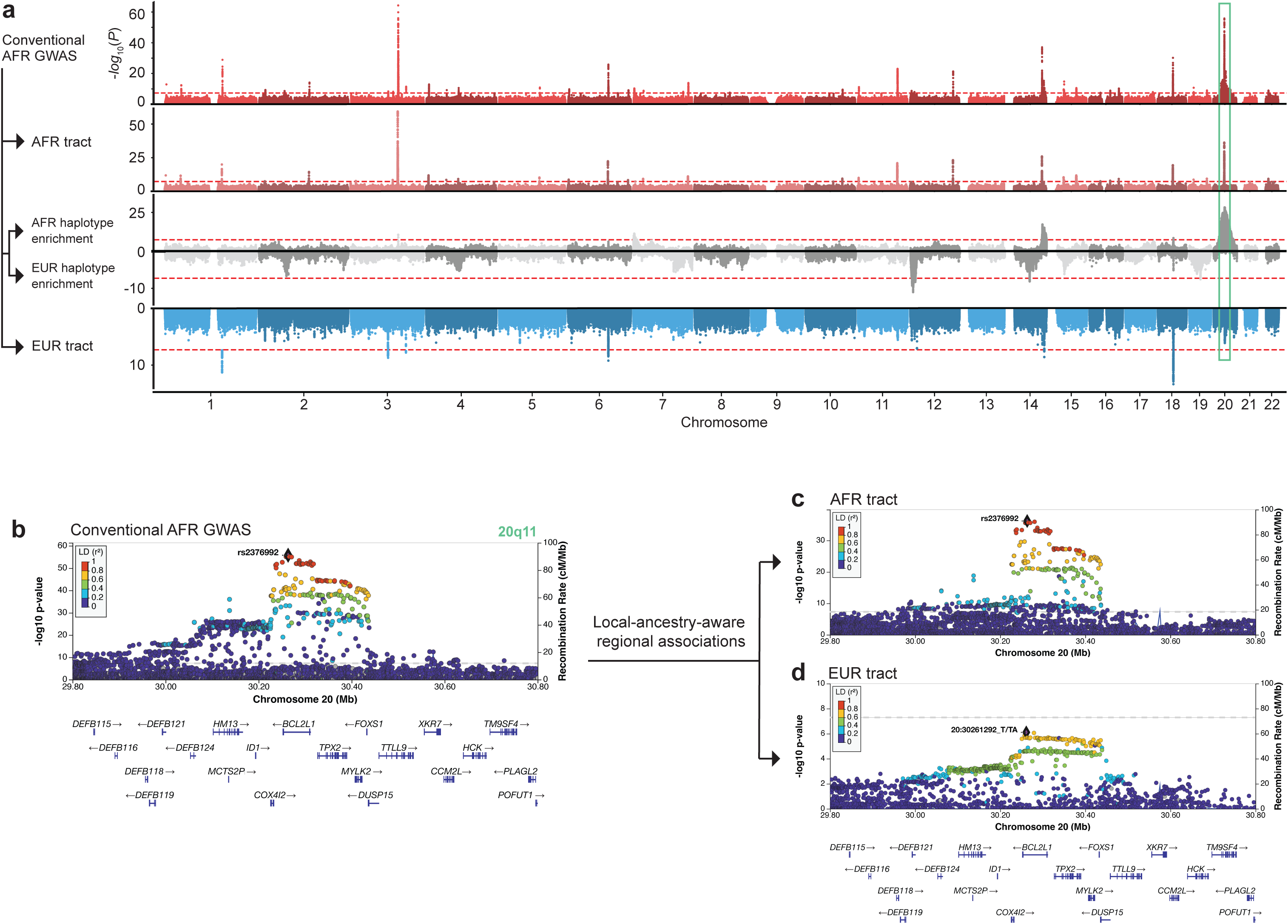
Local-ancestry-aware association analysis of admixed African (AFR) ancestry Million Veteran Program participants. **a**, Stacked Miami plot showing the −log_10_(*P*) values for components of the local-ancestry-aware mLOY GWAS for MVP AFR (from top to bottom): conventional AFR GWAS, AFR tract, AFR haplotype dosage enrichment, European (EUR) haplotype dosage enrichment, and EUR tract. The 20q11 locus is highlighted and shown in the regional Manhattan plots below. **b**, At 20q11 (*BCL2L1*) we observed inflation due to local ancestry. This inflation was resolved by separating the AFR and EUR tracts as shown in **c** and **d**.

Inflation due to admixture was observed at 20q11.21 near *BCL2L1* (Fig. 3ab). Global adjustment for ancestry by including 20 principal components (PCs) in our conventional GWAS model did not sufficiently resolve this locus. We therefore inferred that the inflation was due to the differences in risk allele frequency at the lead SNP (rs2376992) between ancestral haplotypes (51% AFR vs 22% EUR; Supplementary Fig. 10a) at this large effect size locus (OR=0.80 [0.78, 0.82]; *P*=1.7×10^−56^). This was further supported by 20q11.21 having a highly significant difference in AFR and EUR haplotype dosages (*P*=5.6×10^−29^; Fig 2a). In the local-ancestry-aware model, the secondary cluster of significant variants downstream of *BCL2L1* was resolved in the AFR tract relative to the EUR tract (Fig. 3cd; Supplementary Fig. 10b). Strikingly, a meta-analysis of the AFR and EUR tracts for chromosome 20 yielded an inflation factor (λ) of 1.08, compared to 1.40 from the conventional AFR GWAS for this chromosome (Supplementary Data 26). By fine-mapping the smaller credible set of SNPs identified in the AFR tract, we found that the lead risk allele rs2376992 also had the highest posterior probability, and is found in a known promoter region for *BCL2L1* (ENSR00001234227).

At 18q12.3 (*SETBP1*), a known EUR mLOY locus, we observed a complex LD structure with multiple causal SNPs which inhibited fine mapping of this locus in AFR (Supplementary Fig. 11c). Local-ancestry-aware association analysis resolved the overlapping LD structures, and revealed the primary AFR signal at rs4414576 (Supplementary Fig. 11d); this allele has 32% frequency in AFR and only 3% in EUR. The EUR tract at this locus (Supplementary Fig. 11e) has a similar structure to the EUR GWAS (Supplementary Fig. 11f).

Additionally, we performed local-ancestry-aware GWAS in HIS (Extended Data Fig. 6). In this model we assessed the haplotype enrichment of the two main ancestral contributors, EUR and Native American (NAT). Similar to AFR, we found the most significant differential enrichment of haplotypes at 20q11.21, and the same lead variant (rs2376992). This locus in HIS is inflated by the low frequency of rs2376992 in NAT haplotypes (0.3%, compared to 51% in AFR and 22% in EUR).

### Gene set, tissue and cell-type enrichment of mLOY genes

Using *P*-values from the MVP+UKB meta-analysis, we performed MAGMA gene-property tissue expression analysis; this indicated strong specificity for blood (*P*=2.57×10^−16^) and spleen (*P*=1.04×10^−9^) tissues, consistent with our measurement of mLOY in leukocytes (Supplementary Fig. 12a). The majority of independent GWS signals and variants in LD were annotated as intronic (proportion=0.52; Supplementary Fig 12b). Genes mapped within 10Kb of GWS LD blocks were sent to gene set enrichment analysis (GSEA; Supplementary Data 27). The most significantly enriched Kyoto Encyclopedia of Genes and Genomes (KEGG) pathways were p53 signaling, cell cycle, and chronic myeloid leukemia, which reflect key biological processes proximal to mLOY. Hallmark gene sets, representing well-defined biological processes, were used to categorize known and novel mLOY loci (Supplementary Fig. 12c). Genes associated with the G2/M checkpoint were the most significantly enriched with 14 known and 20 novel loci (adj*P*=3.16×10^−6^), followed by genes encoding cell cycle related targets of E2F transcription factors, and genes mediating programmed cell death (apoptosis).

We then tested for enrichment of MVP EUR index variants located in cell-specific open chromatin regions, by intersecting our genetic associations with data from two catalogs of the human epigenome that profile major human body lineages and blood cell lines^36,37^. At the tissue level, we found significant enrichment only in myeloid/erythroid cells (Supplementary Fig. 13a; adj*P*=1.2×10^−4^). Of the blood cell lines, the highest enrichment was measured for multipotent progenitors (MPP; adj*P*=6.4×10^−4^) and their subsequent differentiation stages, i.e. common myeloid progenitors (CMP; adj*P*=1.2×10^−3^) and lymphoid-primed multipotent progenitors (Supplementary Fig. 13b; LMP; BH-corrected *P*=1.1×10^−3^), thus supporting the established role of mLOY genetic effects on blood cell differentiation^21^. Interestingly, among the six differentiated cell types encompassing myeloid, erythroid, and lymphoid cells (Supplementary Fig. 13b), only erythroid cells exhibited significant enrichment (adj*P*=0.017). This pattern of mLOY genetic variant enrichment in differentiating blood cells differs markedly from enrichment patterns observed for other diseases characterized by perturbations in immune responses, such as Crohn’s disease, rheumatoid arthritis, systemic lupus erythematosus, multiple sclerosis or Alzheimer’s disease (Supplementary Fig. 13c).

### eQTLs from single-cell RNA-seq

To identify how SNPs associated with mLOY in EUR associate with gene expression in immune cells, we compared our GWAS signals with a recently published expression quantitative trait loci (eQTL) dataset derived from single-cell RNA-seq data across immune cell populations in Northern Europeans^38^. Across the fourteen immune cell subsets, we found that 197 of 327 MVP EUR mLOY signals spanning 251 eQTLs reached at least nominal significance (*P*<0.05; Supplementary Data 28). Of these, 34 eQTLs (22 unique genes; 8 unique SNPs) reached the significance threshold corrected for multiple testing (*P*<1.1×10^−5^); 20 of these eQTLs were associated with two SNPs (6:29842409:T:C, 6:31150435:A:G) in the major histocompatibility complex (MHC) region (6p22.1 to 6p21.3).

### Multi-tissue TWAS and SMR

We linked our MVP EUR GWAS signals with functional gene units in a multi-tissue transcriptome-wide association study (TWAS). Our TWAS leveraged 43 tissue models including STARNET blood^39^ and a high-powered dorsolateral prefrontal cortex (DLPFC) dataset^40^ (Supplementary Data 29); this yielded 2,297 unique significant gene features at *P_Bonferroni_*<0.05. In the STARNET blood model, 117 features were significant at *P_Bonferroni_*<0.05 (Supplementary Data 30, Supplementary Fig. 14a). In the DLPFC model, 191 features were significant at *P_Bonferroni_*<0.05; revealing one novel gene, *IL21R* (cytokine receptor for interleukin 21) (Supplementary Data 31). All tissues were then meta-analyzed using ACAT^41^, yielding a total of 683 genes with *P_Bonferroni_*<0.05 (Supplementary Data 32, Supplementary Fig. 14b). ACAT revealed an additional four novel genes: *PSTPIP2*, *CCNK*, *PARP10*, and *G3BP1.* We found that mLOY-associated gene expression was highly correlated across the imputed transcriptomes of all tissues (Supplementary Fig. 15), similar to what has been previously observed for other traits^42,43^.

We further performed summary-data-based Mendelian randomization (SMR) experiments to provide support for inference of causality. Across 33 tissue types, we identified 1,870 significant genes with *FDR*<0.05 and *P_HEIDI_*≥0.05, and of these, 234 were identified in blood SMR for MVP EUR (Supplementary Data 33). SMR in blood provided causal support for 23 significant genes (20%) from STARNET blood, and 51 genes (7%) across the combined TWAS findings from STARNET blood, DLPFC, and ACAT meta-analysis (Supplementary Fig. 16); The most significant of these 51 genes from TWAS with causal support include *ATM*, whose gene product repairs DNA double-strand breaks, *NFE2*, a transcription factor that plays a crucial role in hematopoietic differentiation, and *SLC22A5*, whose gene product transports carnitine (required to move fatty acids into mitochondria) into cells (Supplementary Data 33). Of the novel TWAS genes listed above, *PARP10*, whose gene product regulates transcription and DNA repair, had causal support from SMR in blood.

### Genetic correlations of mLOY

Genetic correlations (*r*_g_) between mLOY in MVP EUR and 750 traits were tested (Supplementary Data 34). At the multiple-testing-corrected threshold (α=6.67×10^−5^), 36 traits were significantly correlated. Several metabolite measures had a significant negative *r*_g_ with mLOY, including particle sizes of cholesterol, phospholipids, triglycerides, and total lipids in VLDL, which had *r*_g_ ranging from −0.34 (standard error = 0.07) to −0.28 (0.06). Significant and negative *r*_g_ were observed for hip circumference, and the related anthropometric measures obesity class 2 and BMI. We also generated *r*_g_ using UKB^1^ summary statistics, and found highly similar results as in MVP, except among a small group of obesity-related traits, which had a stronger negative *r*_g_ in MVP (Supplementary Data 34).

### Association of PGS Catalog-based polygenic scores with mLOY

We calculated polygenic scores (PGS) for every trait in the PGS Catalog^44^, for all EUR subjects, and performed a phenome-wide scan for the association of normalized PGS scores with mLOY case-control status (PGS-WAS) to discover shared genetic etiology (Supplementary Data 35). A total of 2,644 scores corresponding to 562 uniquely mapped traits in the Experimental Factor Ontology were tested; we found 82 mapped traits significant after multiple testing correction (α=0.05/562). Many of the most significant traits were blood measures, including increased OR (associated with a standard deviation increase in the PGS) of plateletcrit (1.11 [1.10,1.12]), leukocyte count (1.07 [1.06,1.08]), monocyte count (1.07 [1.06,1.08]), and neutrophil count (1.06 [1.05,1.07]), and decreased mean corpuscular hemoglobin concentration (0.94 [0.93,0.95]). Several metabolic traits were also associated, such as triglycerides (0.93 [0.91,0.94]), HDL (1.06 [1.04,1.07]), BMI (0.95 [0.94,0.96]), SHBG (1.05 [1.04,1.06]), T2D (0.93 [0.91,0.94]), as well as smoking status (1.05 [1.04,1.06]).

### Multi-trait conditioning (mtCOJO) of mLOY on cigarettes per day

In MVP participants, we observed a strong observational association of mLOY with smoking, as well as in the PGS-WAS. To parse out the residual effects of smoking on mLOY susceptibility, we conducted multi-trait-based conditional and joint association analysis^45^ (mtCOJO) in EUR, conditioning on cigarettes per day^46^. Across the genome, only the 15q25 region was primarily impacted by the conditional analysis (Extended Data Fig. 7a). This signal, which was GWS for mLOY in the primary GWAS, was attenuated towards the null after conditioning (Extended Data Fig. 7bc). This locus contains a well-known cluster of smoking-related genes, including *CHRNA5* which encodes a nicotinic acetylcholine receptor subunit that has been frequently associated with smoking and lung cancer in GWAS^47,48^. The *CHRNA5* locus did not reach GWS in MVP AFR nor HIS cohorts. However, the top SNP in the MVP multi-ancestry meta-analysis (rs11852372) reached nominal significance in AFR (*P*=0.03; OR=1.05 [1.01, 1.09]), with a similar effect as observed in EUR (OR=1.04 [1.02, 1.05]).

### Polygenic risk and BMI influence penetrance of mLOY in an age-dependent manner

We observed significant negative associations between mLOY and obesity in PheWAS and PGS-WAS, as well as a negative genetic correlation; this phenotypic association has also been previously reported^12,49^. Therefore, we sought to examine the prevalence of mLOY in MVP EUR as a function of age, BMI, and mLOY PRS decile derived from UKB^1^. In all age bins, we observed that decreasing BMI and increasing PRS were associated with higher mLOY prevalence (Fig. 4a). Our results suggest that the association between mLOY and BMI is not merely confounded by the strong age-dependence of mLOY. We also observed a stronger genetic correlation between mLOY and BMI in MVP EUR (*r*_g_=-0.11 (0.03)) than in UKB (*r*_g_=-0.05 (0.03); Supplementary Data 34); this may partly be a result of ‘healthy volunteer bias’ in UKB^50^.

**Fig. 4.**
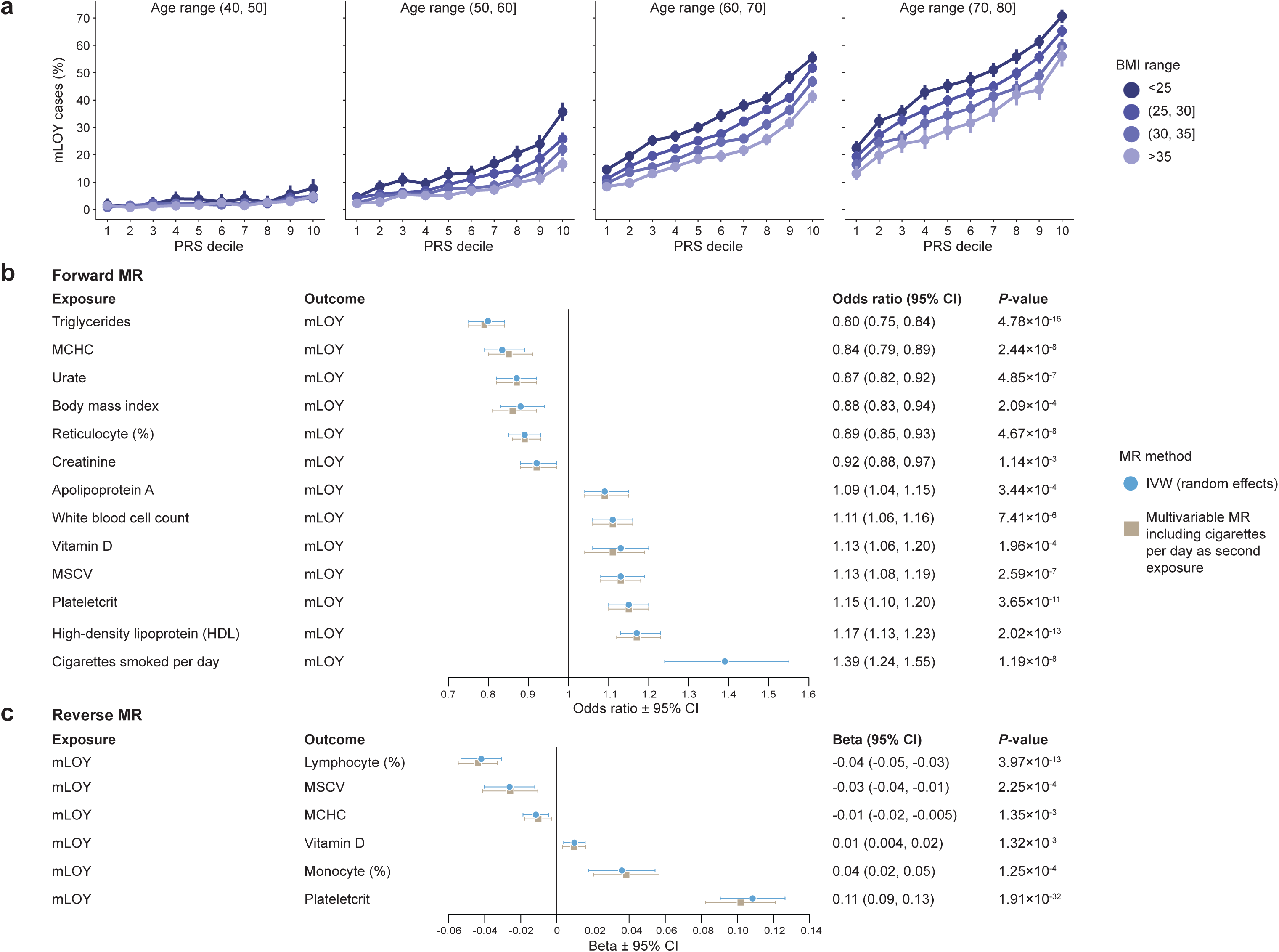
Mosaic loss of Y by PRS decile and Mendelian randomization (MR) **a**, Percentage of individuals with mLOY by PRS decile, stratified by age decile (plot grid) and BMI range (color). **b**, Forest plot showing exposure traits with significant results from random effects inverse-variance weighted (RE IVW) MR and corresponding multivariable MR (MVMR) with cigarettes per day as an additional exposure, on mLOY outcome in Million Veteran Program Europeans. **c**, Significant results from RE IVW MR and MVMR considering mLOY exposure on trait outcomes. MCHC, mean corpuscular hemoglobin concentration; MSCV, mean sphered cell volume; CI, confidence interval.

### Univariable and multivariable Mendelian randomization

We used the significant PGS-WAS associations to inform the selection of 32 traits for two-sample MR in the MVP EUR cohort, using male-only summary statistics where available, to infer potential causality between these exposures and mLOY. Bidirectional causality was assessed, using mLOY as the outcome (“forward MR”) and exposure (“reverse MR”). Random effects inverse-variance weighted (IVW) forward MR supported thirteen traits as potential causal influences on mLOY (*P*<0.05/32; Fig. 4b; Supplementary Fig. 17; Supplementary Data 36). Our analysis indicates that triglycerides, mean corpuscular hemoglobin concentration (MCHC), urate, BMI, reticulocyte percentage, and creatinine exert negative causal influence on mLOY, while cigarettes per day, high-density lipoprotein (HDL), plateletcrit, mean sphered cell volume (MSCV), vitamin D, white blood cell count, and apolipoprotein A (ApoA) showed positive causal influence. Reverse MR supported potential causal influence of mLOY on six traits; indicating negative causal influence upon lymphocyte percentage, MSCV, and MCHC, and positive causal influence upon plateletcrit, vitamin D, and monocyte percentage (Fig. 4c; Supplementary Fig. 18; Supplementary Data 37). All MR results remained robust to multiple sensitivity analyses; all tests except ApoA had *P*<0.05 in weighted median MR^51^, and all tests passed MR-Egger^52^ and MR-PRESSO^53^ tests for evidence of horizontal pleiotropy. The bidirectional MR associations with plateletcrit, vitamin D, and MSCV suggest a shared heritability may underlie those associations.

Because mLOY is strongly associated with smoking, and because the pleiotropic effects of tobacco smoking instruments have large effects on human health, we conducted multivariable MR using cigarettes per day^54^ as a second exposure in the models of significant forward and reverse MR associations (Fig. 4bc; Supplementary Data 38). The direct effect of each exposure on their respective outcomes in multivariable MR were highly similar to the univariable model, and remained significant, with the exception of Vitamin D (forward and reverse) and MCHC (reverse) dropping marginally below the multiple testing threshold. Overall, multivariable MR demonstrates that the inferred causality identified in univariable MR was independent of cigarette smoking. Finally, we sent the significant forward MR traits to mtCOJO and observed modest conditioning of mLOY by triglycerides, particularly at *BUD13* (Supplementary Fig. 19).

## Discussion

In this study we have more than doubled the number of loci associated with mLOY, adding 240 novel risk loci which predispose individuals to this highly polygenic acquired trait. Our discovery was powered by a large cohort of older males within MVP^24^ (N_total_=544,112; N_cases_=126,108), which is further enriched for mLOY risk factors such as cigarette smoking (Supplementary Data 1). This GWAS of mLOY also includes the largest admixed AFR and HIS cohorts to date, for whom we have detailed overlapping genetic risk factors as well as ancestry-specific loci. An additional benefit of MVP is that the extensive EHRs within this biobank enabled a thorough exploration of the associations of mLOY with other phenotypes.

Even with the addition of new mLOY risk loci, our top GSEA results (Supplementary Fig. 12c, Supplementary Data 27) strongly align with the mechanistic hypotheses of earlier GWAS, namely that disruptions in cell cycle regulation and apoptosis are key germline risk factors for mLOY^1,20,211,15,20^. Our most significantly enriched gene set was the G2/M checkpoint (14 known and 20 novel loci), responsible for blocking damaged and incompletely replicated DNA from progressing through the cell cycle. The next most highly enriched gene set, E2F transcription factors, are regulators of the cell cycle^55^, DNA replication, and apoptosis. Apoptosis was the third most highly enriched gene set, and our findings strengthen previous inferences regarding the crucial role of programmed cell death in limiting expansion of mosaic cell populations^20^.

Forward MR analyses revealed several significant blood lipid traits, with triglycerides exhibiting the most significant inferred causal effect on mLOY risk. Triglycerides also had a negative genetic correlation and PGS association with mLOY that were among the strongest we observed, and this trait is also closely linked with the top PheWAS negative associations, obesity and T2D. These relationships can potentially be explained by the ability of triglycerides to induce apoptosis in monocytes^56–58^, a process that can weaken innate immunity but also limit the expansion of clonal cell populations. A similar mechanism may explain the negative causal influence of urate upon mLOY, and the corresponding negative PheWAS and PGS associations with gout, as urate induces apoptosis in monocytes both directly and via reactive oxygen species production^59^. In contrast, the positive causal relationship and PGS association of HDL may be explained by the anti-apoptotic, cytoprotective effects it exerts upon monocytes^60^.

Our significant biomarker traits from forward MR (triglycerides, urate, creatinine, HDL, vitamin D, and ApoA) all replicated the findings from a recent UK Biobank study^61^. That study also highlighted sex hormone-binding globulin (SHBG) as having the strongest inferred causal relationship with mLOY, using a fixed effects IVW model. This relationship was not replicable using a RE model, which is more robust to model assumptions and heterogeneity; in fact, SHBG had among the highest heterogeneity *P*-value of all the traits we tested (*P*_Q_=7.6×10^−130^).

Our study was not without limitations. First, we performed a cross-sectional study at a single time point. Future studies may benefit from a prospective study design, as mLOY is highly associated with age^62^. We also did not consider the effects of environmental exposures aside from smoking and BMI. Veterans may be disproportionately exposed to mutagens over their lifetimes compared to the general public^63^, and these environmental effects may add to or interact with inherited mLOY risk. Next, the majority of mLOY studies, including ours, are based on DNA from peripheral blood mononuclear cells only, limiting our view of mLOY in other disease-relevant tissues. Finally, potential causal relationships inferred by MR should be interpreted cautiously given the high polygenicity of mLOY and potential for residual pleiotropy among genetic instruments.

In summary, we report the largest genomic and phenotypic association scans of mLOY, to date. We identified common genetic architecture across multiple MVP ancestral populations, as well as with the previous UKB^1^ and BBJ^21^ cohorts, strengthening the generalizability of our findings. We hope the novel risk loci identified in this study can enable future researchers to develop improved genetic risk prediction, diagnosis, and understanding of the cellular mechanisms surrounding mLOY.

## Methods

### Ethics/study approval

The VA Central Institutional Review Board (IRB) approved the study protocol. All participants provided informed consent, and all studies conducted at participating centers received approval from IRBs, in accordance with the Declaration of Helsinki. Only previously generated data were analyzed in this study.

### Genotyping, imputation, and ancestry assignment

Genomic data processing was performed for >650,000 MVP participants (releases 1-4). Genotyping was performed using the Thermo Fisher MVP 1.0 Affymetrix Axiom Biobank array^28^. Sample exclusion criteria included: >2.5% missing genotype calls, excess heterozygosity, potential duplicates, and discordance between genetic sex and self-identified gender. SNPs with missingness >5% or MAF that deviated by >10% from the 1000 Genomes Project Phase 3 (1KGP3) data^64^ were excluded. Ancestry was defined using a machine learning algorithm that harmonizes self-reported ethnicity and genetic ancestry (HARE)^65^. Using HARE, we assigned 544,112 male participants in MVP to either European (EUR), African (AFR), or Hispanic (HIS) ancestry.

Genotypes were statistically phased over the entire cohort using SHAPEIT4^66^ (v4.1.3) with Positional Burrow Wheeler Transform (PBWT) depth set to 8. AFR phased genotypes were imputed to the African Genome Resources (AGR) reference panel^67^ using Minimac 4^68^ (v4.1.0). The AGR panel consists of all 5,008 1KGP3 haplotypes and an additional 2,862 haplotypes from unrelated pan-African samples. As AGR contains biallelic SNPs only, a second imputation was performed using 1KGP3 to include indels and other complex variants merged into the primary imputation. Imputation for chrX was performed using the TOPMed panel (hg38); these data became available to us while we were performing this study, and yielded a pronounced improvement in imputation quality for chrX variants. Significant mLOY loci on chrX were lifted over^69^ to hg19 and reported with both build positions.

### Detection of mLOY using long-range haplotype phase

We used SHAPEIT4^66^ to infer haplotypes from array genotypes for the whole MVP cohort and we utilized MoChA^8,25^ (v20200825), an extension to the BCFtools software suite^70,71^, to infer the presence of mLOY by detecting shifts in allelic ratios between phased PAR1 and PAR2 haplotypes, similar to what was done previously in UKB^1^. Briefly, we selected XY males who had chrX mCA calls with length >1 Mb and lod_baf_phase > 2 as mLOY cases. Samples with chrX calls with 1 ≤ lod_baf_phase ≤ 2 were empirically determined to have borderline evidence of mLOY and were not included as cases or controls. This methodology allowed us to infer the presence of mLOY for CFs as low as ∼1%. CF was estimated from B allele frequency deviation (bdev) using the formula 4*bdev / (1+2*bdev).

### Estimating effect of ancestry and smoking status

We estimated the effects of ancestry and smoking status on mLOY. Effects of AFR and HIS ancestries were estimated with reference to EUR. Smoking status (current, former, never) was ascertained from smoking-related health factors and ICD-9/10 codes for tobacco dependence, using an algorithm previously validated in the VA^72^. Covariate sensitivity models included age, age-squared, BMI, and ancestry or smoking status where applicable and logistic regression was performed in R (v4.2.2). Because of the imbalance of covariate distribution, particularly age, between MVP ancestries, we generated balancing weights for participants as an additional effects estimation sensitivity analysis. We used WeightIt^73^ (v1.3.0) with the average treatment effect in the population (ATE) estimand and two weighting methods, generalized linear model (glm) propensity score weighting and entropy balancing. Marginal odds ratios were generated with g-computation using marginaleffects^74^ (v0.23.0) for R.

### Phenome-wide association studies

ICD-9/10 codes were grouped hierarchically into 1,605 phecode-based disease traits. For each phecode, individuals with two or more relevant codes were set as cases, and those without codes were considered controls. Associations of phecode-based outcomes on mLOY status were tested within HARE groups using the R PheWAS package^75^ (v0.1). Age, age-squared, and the first 20 ancestry-specific PCs were included as covariates. A Bonferroni-corrected threshold was used to assess significance (α=0.05/1605).

### GWAS

Presence or absence of any detectable mLOY CF as determined by MoChA^8,25^ was used as the case-control designation in all GWAS. Single variant genome-wide association testing was carried out with REGENIE^76^ (v1.0.6.7) using age, age-squared, and the first 20 ancestry-specific PCs. REGENIE applies a whole-genome regression model to control for relatedness and population structure, and includes a Firth correction to control for bias in rare SNPs. REGENIE step 1 was performed using leave-one out cross validation (--loocv). Approximate Firth likelihood ratio test (LRT) was applied as fallback for associations with *P*<0.05, with SE computed based on LRT where applied (--firth-se). We kept common variants with MAF≥0.1% and imputation quality (INFO) >0.3. A higher INFO cutoff of >0.5 was used for chrX, due to greater challenges with X chromosome genotyping and imputation, and availability of fewer reference haplotypes due to male haploidy. GWAS Manhattan plots were generated using gwaslab^77^ (v3.4.7).

Two significant chrX loci in PAR1 (rs2857319, *XG*) and PAR2 (rs306890, *SPRY3*), both near the boundary with the nonPAR, had large frequency differences between X and Y chromosomes in the Genome Aggregation Database (gnomAD v3.1.2). These loci were removed after determining the mLOY-associated allele was exclusively in high CF participants, indicating likely genotyping error (Supplementary Fig. 20).

Within each ancestry group, we performed conditional association analyses using Genome-wide Complex Trait Analysis multi-SNP-based conditional and joint association analysis (GCTA-COJO)^78^ to identify secondary association signals at associated loci, using LD reference panels consisting of 100,000 randomly selected participants for EUR and AFR, and all 52,183 MVP participants in HIS. Default values for all parameters were used. COJO output SNPs with *r*^2^ 0.05 were iteratively retained based on lowest *P*-value.

For replication, MVP EUR COJO association signals were compared to summary statistics from the previous mLOY GWAS in UKB^1^. An updated version of UKB chrX summary statistics were utilized for this comparison (N_case_=40,466; N_control_=146,066). These were run natively on GRCh38 using the pipelines ukb2txt.wdl, mocha.wdl, impute.wdl, and assoc.wdl from https://github.com/freeseek/mochawdl. MACH R2 values were considered for variant quality in lieu of INFO scores. The updated chrX sumstats contained an additional signal which did not previously reach GWS^1^, X:1565178:C:T (hg38).

### GWAS sensitivity analysis in high CF mLOY cases

We performed an additional GWAS sensitivity analysis in only mLOY cases with high CF (>10%). GWAS methods, including QC, model parameters, and post-processing with COJO, were conducted identically to the full cohort models described above. Because selecting for high CF cases exacerbated the age difference between cases and controls (>1 standard deviation in AFR and HIS), we matched cases and controls in each ancestral group on age and age-squared before performing this GWAS. Each case was pre-processed to match two controls in EUR, or three controls in AFR and HIS, with nearest-neighbor (greedy) matching without replacement, using MatchIt^79^ (v4.5.5) with matching distance between participants calculated by general linearized model. After matching, absolute standardized mean difference (ASMD) values were <0.05 in AFR and approximately equal to 0.2 in EUR and HIS, indicating improved covariate balance.

### Fine-mapping

We adapted the FinnGen fine-mapping pipeline^80^ (https://github.com/FINNGEN/finemapping-pipeline) to perform Bayesian fine-mapping at each genome-wide significant locus in EUR, AFR, and HIS using Sum of Single Effects (SuSiE) modeling^81^. Pairwise SNP correlations were calculated directly from imputed dosages on 320,831 European-ancestry samples in MVP using LDSTORE 2.0^82^. The maximum number of allowed causal SNPs at each locus was kept at the default of 10. Fine-mapping regions which overlapped the MHC region (chr6:25,000,000-34,000,000) were excluded. High quality credible sets were defined as those with minimum *r*^2^<0.5 between variants (88/422 discarded in EUR, 24/69 in AFR, 3/21 in HIS).

### Rare variant analysis

We conducted association analyses for rare variants in each MVP ancestry group using REGENIE and the same covariates as in standard GWAS. We considered only variants directly genotyped on the MVP 1.0 array^28^, which is enriched in protein-altering rare variants, and applied the Rare Heterozygous Adjustment algorithm^83^ to improve the positive predictive value of rare genotype calls. We further restricted the included markers to directly genotyped ultra-rare variants (MAF<0.1% in controls) classified as “high-impact”^84^. Rare variants were categorized as somatic or germline based on allele balance for heterozygotes obtained from the Genome Aggregation Database (gnomAD)^85^. For a heterozygous germline variant, the distribution of allelic balance in heterozygotes has a peak at 50%, while for somatic variants the allelic balance distribution has a peak closer to 0%, with a wide range of allelic balances from 0-100% representing varying cell fractions.

### GWAS multi-ancestry meta-analysis

For meta-analysis, MVP EUR, AFR, and HIS cohorts were filtered by INFO>0.5 to retain only high quality variants. Similarly, for the combined meta-analysis of MVP+UKB, the three MVP ancestry groups were meta-analyzed together with UKB^1^ summary statistics comprising 41,791 mLOY cases and 163,220 controls. Each set of summary statistics was converted into GWAS-VCFs using the +munge plug-in of bcftools^71^ (v1.21). Fixed-effect meta-analyses were performed using the +metal bcftools plug-in with an IVW scheme. Only variants present in two or more ancestries were retained in meta-analysis results.

Loci were defined using the two-stage “clumping” procedure implemented in the Functional Mapping and Annotation (FUMA) platform^32^ (v1.5.2). In this process, genome-wide significant variants were collapsed into LD blocks (*r*^2^>0.6) and subsequently re-clumped to yield approximately independent (*r*^2^<0.1) signals; adjacent signals separated by <250kb were ligated into a single locus.

Overlapping loci between cohorts were defined using a ±500 Kbp window from COJO index variants, or in the case of meta-analysis loci, the larger of a ±500 Kbp window and the locus boundaries defined by FUMA as described above. Novelty was defined as the absence of previously reported mLOY associations in the larger of either these locus boundaries or a ±1 Mb window. (The ±1 Mb window was also used to assess novelty of COJO index variants and rare variants).

In the MVP multi-ancestry meta-analysis and meta-analysis of MVP+UKB, we further performed a sensitivity analysis using the Han-Eskin random effects model (RE2) in METASOFT^33^ (v2.0.1). FE and RE2 *P*-values at top loci were highly similar. We replicated our meta-analysis lead variants in UKB^1^ as described above and in BBJ^21^ where available. BBJ reported their results as mLRR-Y intensity thresholding as a proxy for mean Y chromosome dosage in circulating blood cells of subjects.

### Local ancestry inference (LAI) and local-ancestry-aware GWAS

To improve interpretation of GWAS results in admixed groups^35^, we generated local ancestry calls in MVP within 125,846 AFR participants assuming two-way (AFR/EUR) admixture, and within 53,820 HIS assuming three-way (AFR/EUR/NAT) admixture; these were later subsetted by those passing GWAS QC described above. RFMIX2^34^ (v2.03) was used to query MVP release 4 phased genotypes using the gnomAD (v3.1.2) Human Genome Diversity Project (HGDP) + 1KGP subset release as reference. Expectation-maximization (EM) optimization was applied using two and three iterations of the AFR and HIS models, respectively. Global ancestry proportion estimates were derived by taking the average per-chromosome Q estimates (weighted by chromosome length) for each component ancestry. HIS admixture was plotted using ggtern^86^ (v2.2.0).

We then extracted ancestry-specific dosages from the imputed data into files compatible with the PLINK 2.0^87^ (v2.00a3) local covariates feature (--local-covar; --local-pvar; --local-psam). For AFR, EUR-specific allele dosages were loaded into a .pgen (PLINK 2.0 binary genotype) file, and African-specific dosages and EUR haplotype counts were interlaced in a zstandard-compressed table. For HIS, EUR-specific dosages were put into a .pgen file, with African and NAT-specific dosages and EUR and AFR haplotype counts interlaced into a zstandard-compressed table. local-ancestry-aware GWAS was then conducted to obtain ancestry-specific marginal effect estimates.

### Gene set, tissue and cell type enrichment analysis

FUMA GENE2FUNC^32^ was run using MVP+UKB meta-analysis summary statistics in genes that were positionally mapped to significant variants (within 10 Kbp) excluding the MHC gene region. Benjamini-Hochberg (FDR) was used as the gene set enrichment multiple test correction method. Hallmark gene sets^88^ were used to categorize genes.

To further evaluate whether the genomic loci implicated in mLOY were enriched in any particular cell type, we intersected common mLOY risk variants with broad and blood-specific epigenomic catalogs of cell-specific open chromatin^36,37^ using the LD score partitioned heritability approach of LDSC^89^ (v1.01). For the broad epigenome catalog encompassing various human tissues^36^, we re-used the open chromatin regions associated with each tissue from the lists provided by the creators of the atlas (https://www.meuleman.org/DHS_Index_and_Vocabulary_hg38_WM20190703.txt.gz). To identify cell-specific chromatin regions within the epigenome map of human blood lineages^37^, we conducted differential analysis on sequencing data sourced from the Gene Expression Omnibus (GEO) under accession GSE74912. To ensure a consistent evaluation of the generated LDSC statistics, which rely on the overall genomic coverage of the tested chromatin regions, we selected an identical number of the most specific open chromatin regions from each blood lineage for subsequent heritability analysis by LDSC. For contextual comparison of heritability signal with the other diseases, we acquired summary statistics of Crohn’s disease^90^, rheumatoid arthritis^91^, systemic lupus erythematosus^92^, multiple sclerosis^93^, or Alzheimer’s disease^94^. Default parameters of LDSC were used and the MHC region was excluded.

### eQTL analysis using published datasets

We interrogated a previously published dataset^38^ for SNPs associated with mLOY in our European ancestry cohort. Chromosome, position, and alleles were used as unique identifiers with which to cross-reference SNPs across different immune cell subsets.

### Transcriptomic imputation model construction and transcriptome-wide association study

Transcriptomic imputation models were constructed as previously described^42,95^ for tissues of the GTEx^96^ (v8), STARNET^39^ and PsychENCODE^40,97^ cohorts. For GTEx and STARNET cohorts, we considered adipose tissue: subcutaneous (GTEx & STARNET) and visceral (GTEx & STARNET); arterial tissue: aorta (GTEx & STARNET), coronary (GTEx), mammary (STARNET), and tibial (GTEx); blood (GTEx & STARNET); cell lines (GTEx): EBV-transformed lymphocytes and transformed fibroblasts; endocrine (GTEx): adrenal gland, pituitary, and thyroid; colon (GTEx): sigmoid and trasverse; esophagus (GTEx): gastroesophageal junction, mucosa and mascularis; pancreas (GTEx); salivary gland minor (GTEx); stomach (GTEx); terminal ileum (GTEx); heart (GTEx): atrial appendage and left ventricle; liver (GTEx & STARNET), skeletal muscle (GTEx & STARNET); nerve tibial (GTEx); reproductive (GTEx): mammary tissue, ovary, prostate, testis, uterus, vagina; lung (GTEx); skin (GTEx): not sun exposed suprapubic and sun exposed lower leg; and spleen (GTEx). From PsychENCODE^40,97^ we considered brain: dorsolateral prefrontal cortex (DLPFC) genes. The genetic datasets of the GTEx^96^, STARNET^39^ and PsychENCODE^97^ cohorts were uniformly processed for quality control (QC) steps before genotype imputation as previously described^42,95^. We restricted our analysis to samples with European ancestry as previously described^42^. Genotypes were imputed using the University of Michigan server^68^ with the Haplotype Reference Consortium (HRC) reference panel^98^. Gene expression information was derived from RNA-seq gene level counts, which were adjusted for known and hidden confounders, followed by quantile normalization. For GTEx, we used publicly available, quality-controlled, gene expression datasets from the GTEx consortium (http://www.gtexportal.org/). RNA-seq data for STARNET were obtained in the form of residualized gene counts from a previously published study^39^. For the dorsolateral prefrontal cortex from PsychENCODE we used post-quality-control RNA-seq data that were fully processed, filtered, normalized, and extensively corrected for all known biological and technical covariates except the diagnosis status^40^ as previously described^95^. Feature types queried include genes, long non-coding RNA (lincRNA), microRNA, processed transcripts, pseudogenes, RNA, small nucleolar RNA (snoRNA), plus constant (C), joining (J), and variable (V) gene segments.

For population classification we used individuals of known ancestry from 1KGP3. We excluded variants in regions of high linkage disequilibrium, variants with MAF<0.05, variant with high missingness (>0.01), and variants with Hardy-Weinberg equilibrium *P*<1×10^−10^; the remaining variants were pruned (--indep-pairwise 1000 10 0.02 with PLINK 2.0^99^) and PCA was performed with PLINK 2.0^87^. We used the first three ancestral PCs to define an ellipsoid based on 1KGP3 (v5) EUR samples^64^ and samples <3 s.d. from the ellipsoid center were classified as EUR; based on this definition of EUR samples, we excluded one non-EUR ancestry individual. In the remaining samples (n=405), we performed additional sample-level quality control by retaining non-related samples (--king-cutoff 0.0884 with PLINK 2.0^87^) with sample-level missingness < 0.015, variants with variant-level missingness < 0.02, and heterozygosity rate of <3 s.d. from the mean; no samples were excluded by these steps. For the next step of our pipeline, we performed outlier testing in the gene expression data. After performing counts per million filtering (>0.5 counts per million in at least 30% of samples) and voom normalization, PCA was performed, and we excluded individuals located >4 s.d. away from the mean of the ellipsoid defined by PC1 to PC3. This did not remove any individuals and assured us that our data did not contain any outliers. In this final set of individuals, we performed variant-level quality control of the genotypes by removing variants with MAF<0.01 (for all variants possible, we utilized MAFs reported by Allele Frequency Aggregator European population^100^, to reduce MAF bias from the comparatively small imputation model training population), 5 minor allele counts and 0.02 missingness rate; only variants present in the HRC reference panel were retained to ensure good representation of variants in the target GWAS^98^. We used this final set of quality-controlled genotypes in conjunction with our normalized expression data to discover the optimal number of PEER factors to find eQTLs. Our analysis led to the decision to utilize 15 PEER factors, which had resulted in the discovery of 4,299 significant eQTLs^101^. This was the closest value to 90% of the maximum value of eQTLs discovered by any chosen number of PEER factors (4,844 significant eQTLs from 50 PEER factors). This allowed us to retain the maximum signal for gene expression prediction without overcorrecting our data. After residualization for 15 PEER factors, expression data were quantile normalized. Genotypes were then converted to dosages, and missing values were replaced with twice the variant’s MAF before dosages were rounded to the nearest whole number. For training, we used PrediXcan^102^ for the construction of the retinal transcriptomic imputation model due to a lack of SNP epigenetic annotation information; for all other models, we used EpiXcan^42^.

### Multi-tissue transcriptome-wide association study (TWAS)

We performed a gene-trait association analysis as previously described^42^. We applied the S-PrediXcan method^103^ to integrate the summary statistics and the transcriptomic imputation models constructed above to obtain gene-level association results. *P*-values were adjusted for multiple testing using the Benjamini & Hochberg (FDR) method and Bonferroni correction. *P*-values across tissues were meta-analyzed using ACAT^41^ ≤0.05 and predictive *r*^2^>0.01 to control for both significance and variance explained.

### Summary-data-based Mendelian randomization

To test for joint associations between GWAS summary statistics SNPs and eQTL, the SMR method^104^, an MR approach, was used. Top SNPs used in SMR for each probe were selected as the most significant SNP in the eQTL data which was also present in the GWAS data. SMR^104^ (v1.03) was run using the default settings using the same GTEx^96^ (v8) tissues as in the TWAS section. European 1KGP3 samples were used as a reference panel. Bonferroni multiple-testing correction was applied on SMR *P*-values (PSMR). A post-filtering step was applied by conducting heterogeneity in dependent instruments (HEIDI) test. The HEIDI test distinguishes the causality and pleiotropy models from the linkage model by considering the pattern of associations using all the SNPs that are significantly associated with gene expression in the cis-eQTL region. The null hypothesis is that a single variant is associated with both trait and gene expression, while the alternative hypothesis (*P*_HEIDI_<0.05) is that trait and gene expression are associated with two distinct variants.

### Heritability and genetic correlation analyses

Genetic correlation analyses were performed using LDSC^105^ (v1.01) using the provided European-ancestry LD scores derived from 1KGP3, as implemented in LDHub (v1.9.0)^106^. Bonferroni multiple testing correction was applied. SNPs from the MHC region were removed.

### Association of PGS Catalog-based polygenic scores with mLOY status

Phenome-wide polygenic score files for 2,652 traits were obtained from European Molecular Biology Laboratory’s European Bioinformatics Institute (EMBL-EBI) PGS Catalog^44^ (September 2022 version). All EUR-ancestry subjects in MVP were scored across all available PGSs, excluding those derived from other MVP studies (20), using the +score plugin of bcftools^71^. PGSs were loaded into the dosage format field of VCFs readable by SAIGE^107^ (v1.1.6.2) for association testing. Logistic regression was used to examine associations of PGSs on MVP EUR mLOY cases and controls, adjusting for the same covariates as in GWAS (age, mean-centered age-squared, and 20 ancestry-specific PCs).

### Multi-trait-based conditional and joint analysis (mtCOJO)

In order to assess the residual effects of genetic predisposition to cigarette smoking from mLOY susceptibility, we conducted a multi-trait-based conditional and joint analysis (mtCOJO)^45^, conditioned on cigarettes per day^46^. The conditional meta-analyses were performed using the MVP EUR mLOY summary statistics. The EUR LD panel described above for use with COJO was also used in this analysis.

### Polygenic risk scoring

UKB^1^ summary statistics were used to construct a polygenic risk score for EUR MVP participants with PRS-CS^108^ (v20210604) with a global shrinkage prior of 1×10^−4^. European samples of the 1KGP were used as a reference panel. Variants were filtered to include only those with *r*^2^>0.8 and MAF>1%.

### Univariable and multivariable Mendelian randomization

Forward and reverse two-sample MR was performed using summary statistics from MVP EUR and publicly available European GWAS. Summary statistics were retrieved through the OpenGWAS database API^109^ via the accession codes listed in **Supplementary Data 36-37**, except BMI in males only which was derived from the GIANT (Genetic Investigation of ANthropometric Traits) consortium study^110^, and cigarettes per day from the GSCAN (GWAS & Sequencing Consortium of Alcohol and Nicotine use) consortium^54^, which were downloaded separately. The GWS threshold of *P*<5×10^−8^ was used for the selection of genetic instrumental variables. LD clumping of *r*^2^<0.001 within a 10 Mb window was used to identify independent instruments. HEIDI filtering of pleiotropic SNP outliers (*P*<0.01) was implemented using the GSMR2^111^ (v1.1.1) R package.

Selection, clumping, and harmonization of instruments was performed using TwoSampleMR^112^ (v0.6.8). Primary analyses used the random-effect IVW method. Sensitivity analyses were performed with the MendelianRandomization^113^ (v0.9.0) R package using fixed effect IVW, using a nominal significance threshold (*P*<0.05) in the weighted median method and *P*>0.05 for MR-Egger^114^ intercept. MR-PRESSO^53^ was conducted using the MRPRESSO (v1.0) R package to test for horizontal pleiotropy. MR-PRESSO was not performed if either the MR-PRESSO global test was insignificant or the number of IVs retained after outlier removal was insufficient. All *P*-values from MR tests were derived from two-sided tests.

To control for the possibility that genetic instruments related to mLOY exhibited possible horizontal pleiotropic effects via smoking behavior, we conducted multivariable MR using the MVMR^115^ (v0.4) R package and included a second exposure of cigarettes per day^54^. We report IVW multivariable MR results along with the test for heterogeneity from a modified form of Cochran’s Q statistic with respect to differences in multivariable MR estimates across the set of instruments. Covariance between the effect of genetic variants derived from the two exposures was fixed to zero due to the use of non-overlapping samples. The *F*-statistic for instrument strength achieved *F*>10 for all tests.

Significant forward MR traits (triglycerides, HDL, BMI, total testosterone, and SHBG) were sent to conditional analysis with GCTA-mtCOJO^45^. mtCOJO was performed using the MVP EUR mLOY summary statistics and EUR LD panel as described above.

**\*Consortium authors and affiliations**

**VA Million Veteran Program**

J. Michael Gaziano^33,34^, Philip S. Tsao^16,17,18^, Saiju Pyarajan^1,32^

^33^Massachusetts Veterans Epidemiology Research and Information Center (MAVERIC), VA Boston Healthcare System, Boston, MA, USA; ^34^Division of Aging, Department of Medicine, Brigham and Women’s Hospital, Harvard Medical School, Boston, MA, USA

## Data availability

The full summary level association data from the meta-analysis and individual population association analyses in MVP will be available via the dbGaP study accession number phs001672. Full transcriptome-wide association study results are available upon request.

## Extended Data Figure Captions

**Extended Data Fig. 1.**
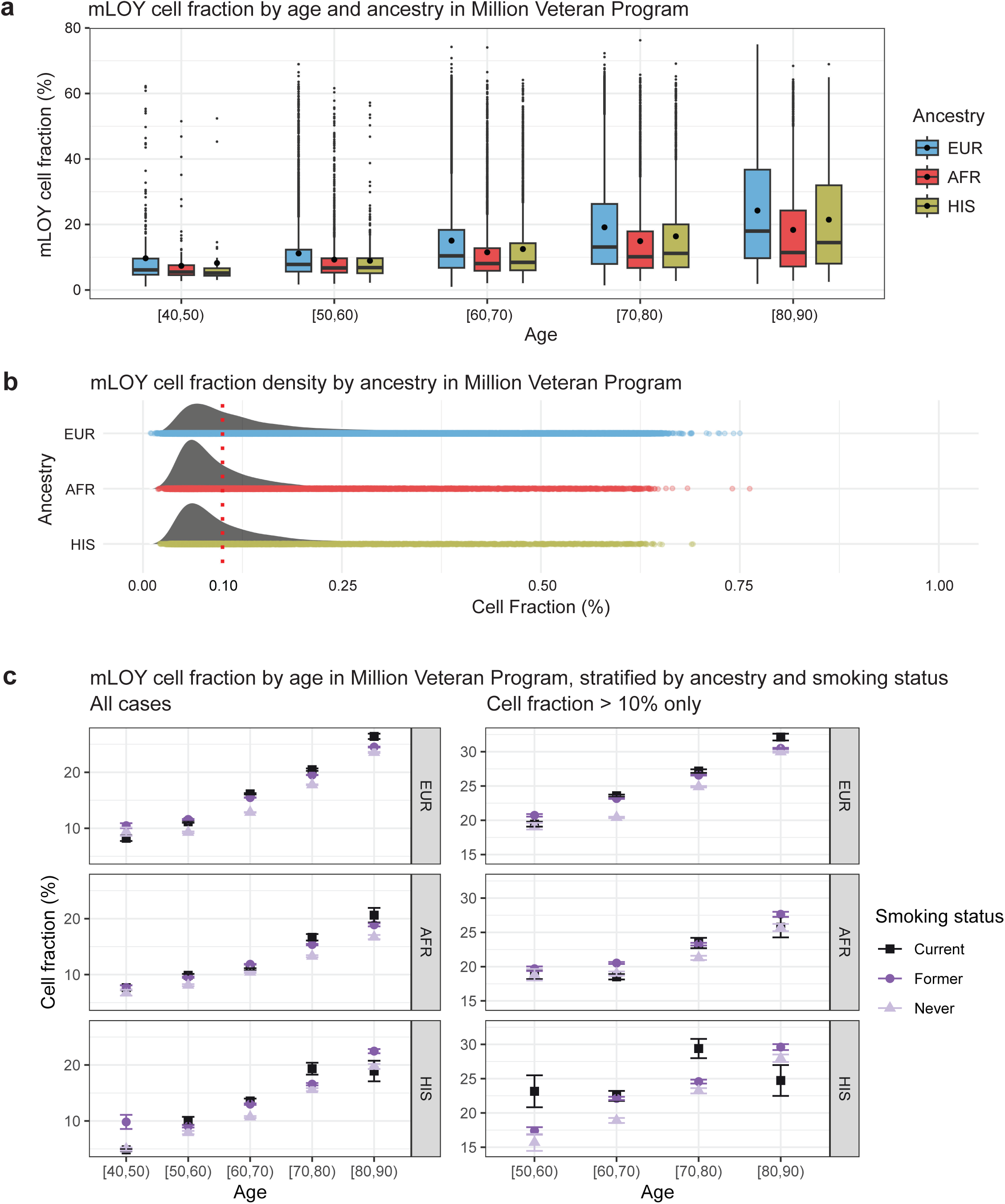
Cell fraction (CF) of mLOY across groups. **a**, CF of mLOY in Million Veteran Program participants by ten-year age bins, stratified by ancestry. Boxplots show first decile, first quartile, median, third quartile, and last decile; dots indicate mean. **b**, Density ridgeline plots show distribution of CF for each ancestry. CF 10% is marked to indicate the cutoff for high CF sensitivity analyses. **c**, CF of mLOY, stratified ancestry and by current, former, and never smoking status. Full cohort and CF>10% are shown. Error bars indicate 95% confidence intervals.

**Extended Data Fig. 2.**
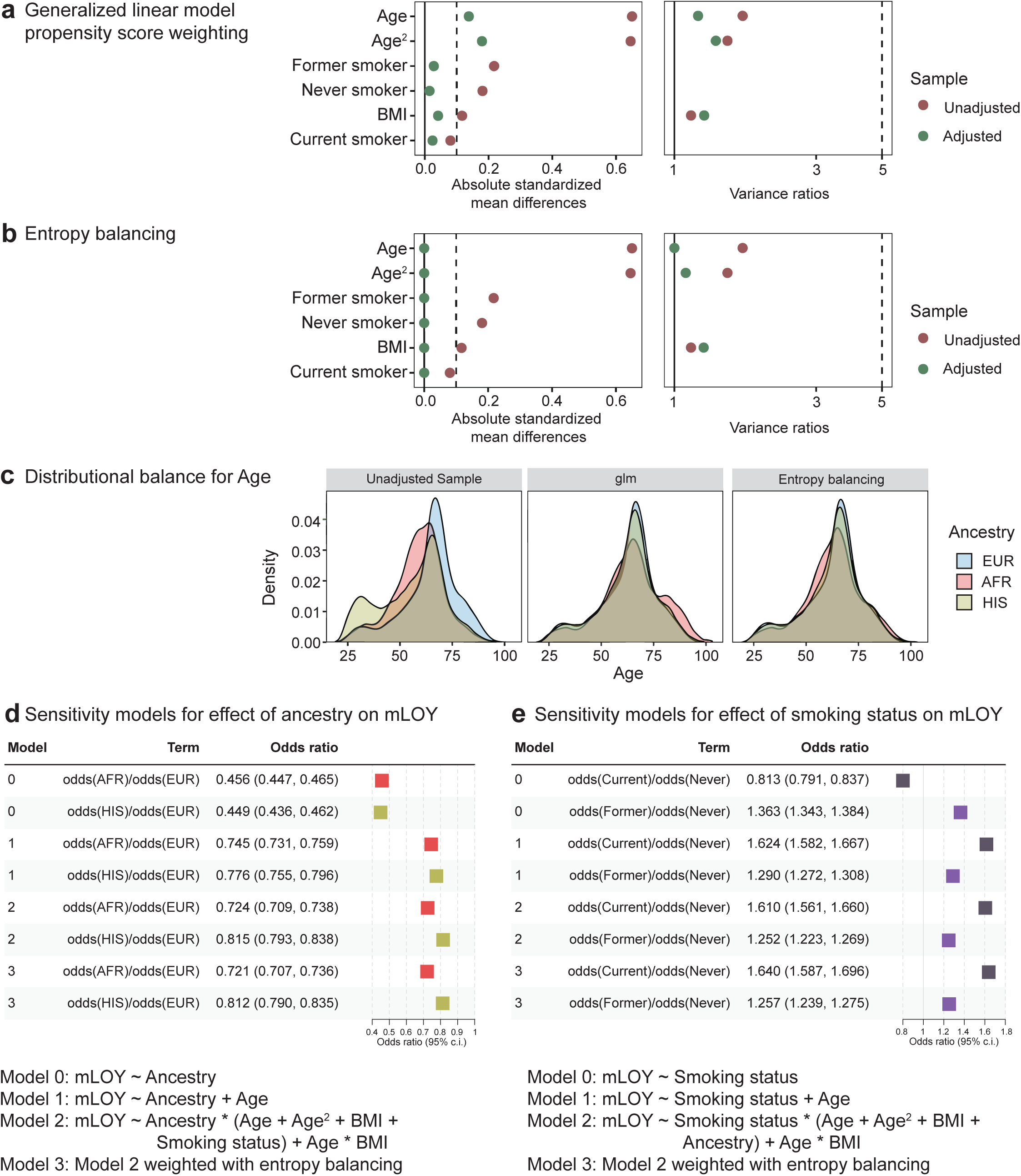
Estimating effects of ancestry and smoking status on mLOY. **a**, Love plot of covariate balance before and after weighting using general linearized model propensity scoring. **b**, Love plot of covariate balance before and after weighting using entropy balancing. **c**, Density plots of age distribution across ancestries before and after each weighting method. **d**, Forest plot of sequential covariate adjustment sensitivity analyses for effect of ancestry on mLOY. **e**, Forest plot of sequential covariate adjustment sensitivity analyses for effect of smoking status on mLOY. BMI, body mass index; c.i., confidence interval. Parenthesis around model 2 covariates indicate the interactions of these terms and ancestry were included in the model.

**Extended Data Fig. 3.**
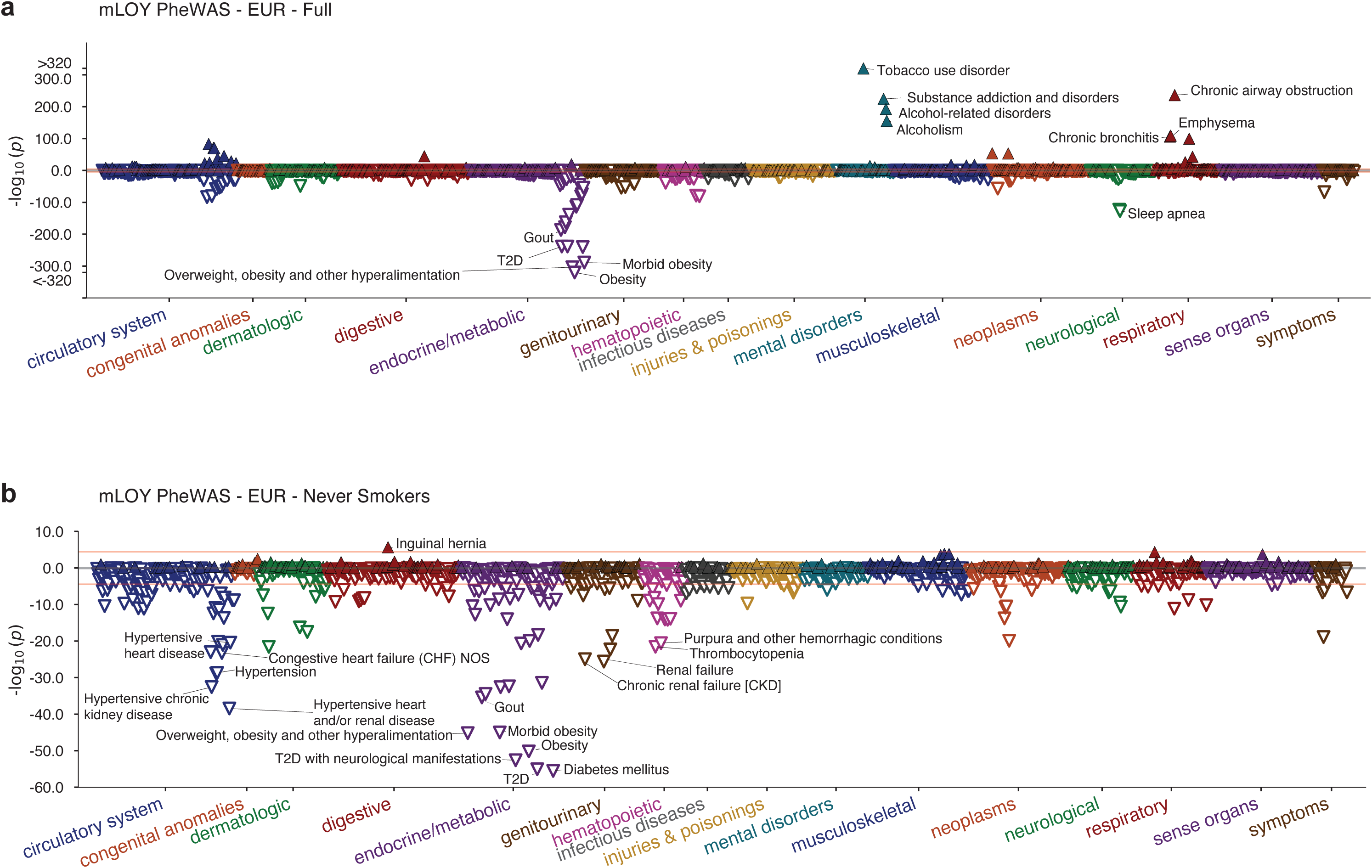
Phenome-wide association studies (PheWAS) of mLOY status. PheWAS for the largest Million Veteran Program ancestry, European (EUR), in: **a**, Full cohort; **b**, Never smokers only. Association *P*-values were obtained from a two-sided test of the z-statistic, calculated using logistic regression. The red line in each plot indicates Bonferroni-corrected significance (α=0.05/1605).

**Extended Data Fig. 4.**
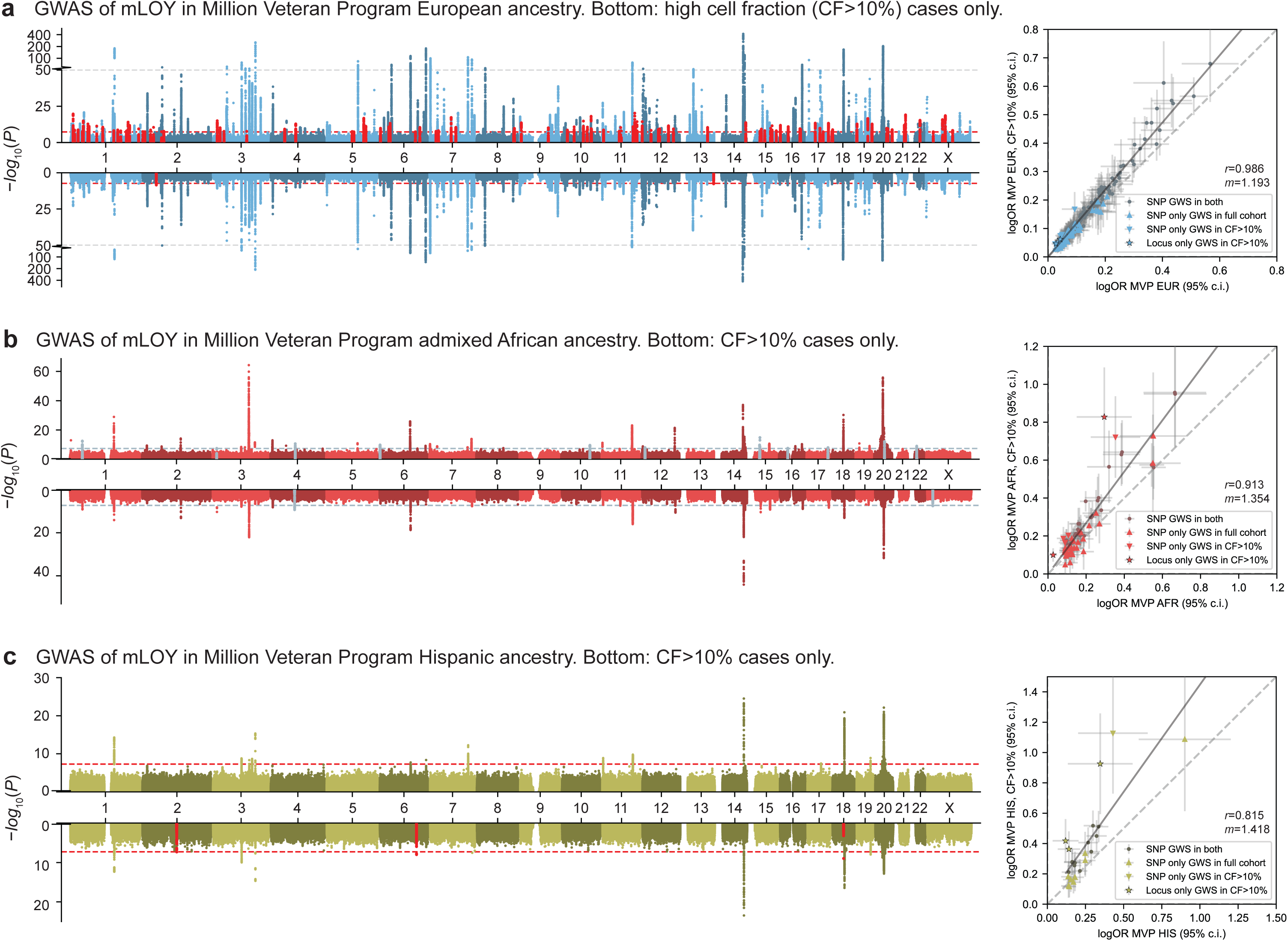
Miami plots of mosaic loss of chromosome Y (mLOY) risk loci in Million Veteran Program participants by ancestry. Miami plots (left) and effect concordance scatterplots (right) for GWAS of mLOY in full cohort and high cell fraction (CF>10%) sensitivity analyses. Miami plots show the −log_10_(*P*) for associations of genetic variants with mLOY in **a**, European ancestry, **b**, African ancestry, and **c**, Hispanic ancestry. Top plots show full cohorts, bottom plots show high cell fraction (CF>10%) sensitivity analyses. Numbers central to plots indicate chromosomes. Novel mLOY index variants (± 50 Kb) are highlighted in top, those additional loci found in CF>10% only are highlighted in bottom plots. The dotted line indicates the genome-wide significance threshold (*P*<5×10^−8^). Light grey dotted lines represent a transition from linear to log-scale on the y-axis. c.i., confidence interval; *r*, Pearson correlation coefficient; *m*, slope.

**Extended Data Fig. 5.**
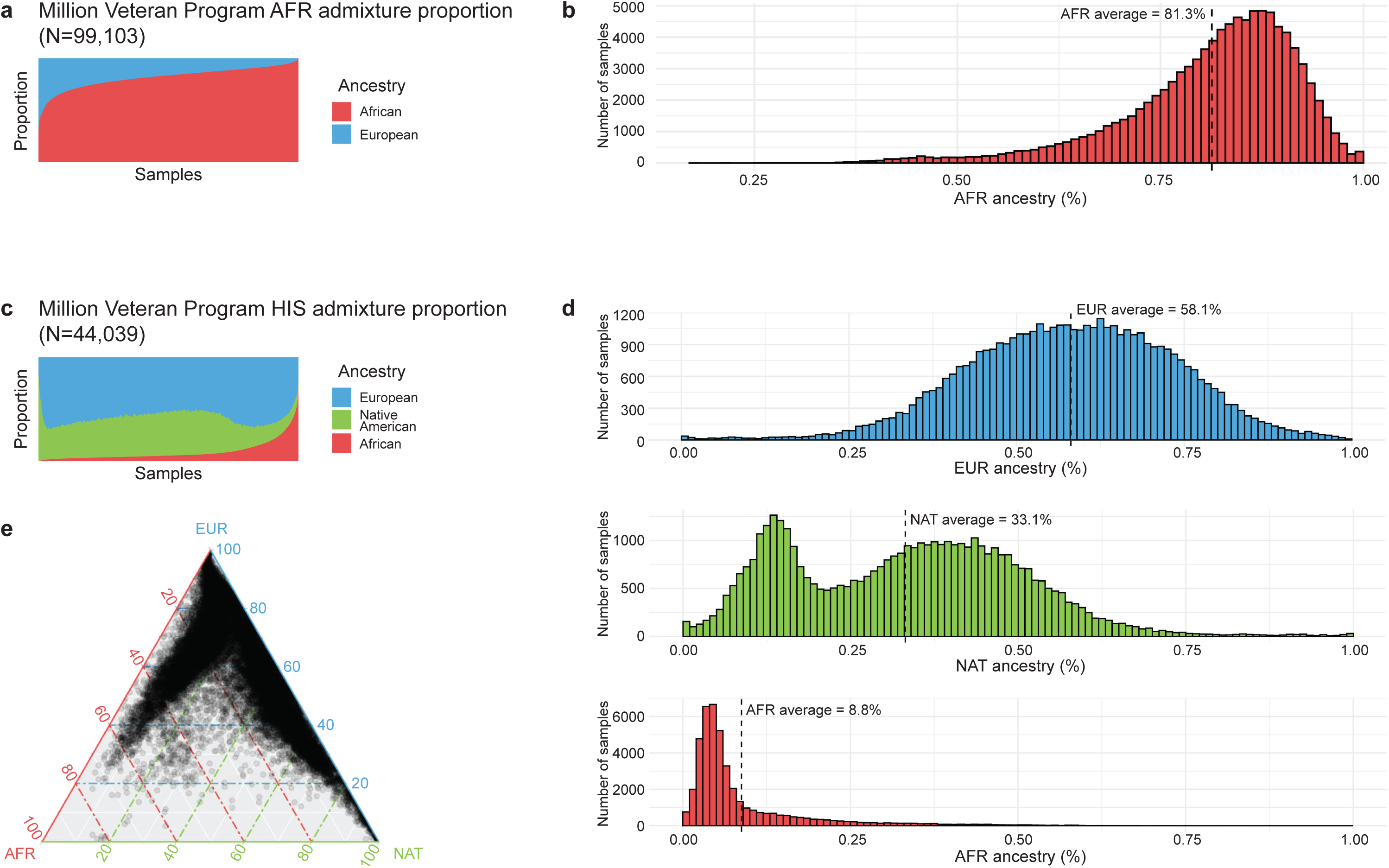
Admixture proportions in Million Veteran Program (MVP) African (AFR) and Hispanic (HIS) cohorts. **a**, Two-way admixture proportion across MVP AFR participants. **b**, MVP AFR admixture binned by percentile of AFR ancestry (%). **c**, Three way admixture proportion across MVP HIS participants. **d**, MVP HIS admixture binned by percentile of European (EUR), AFR, and Native American (NAT) ancestry (%). **e**, Ternary plot of HIS admixture.

**Extended Data Fig. 6.**
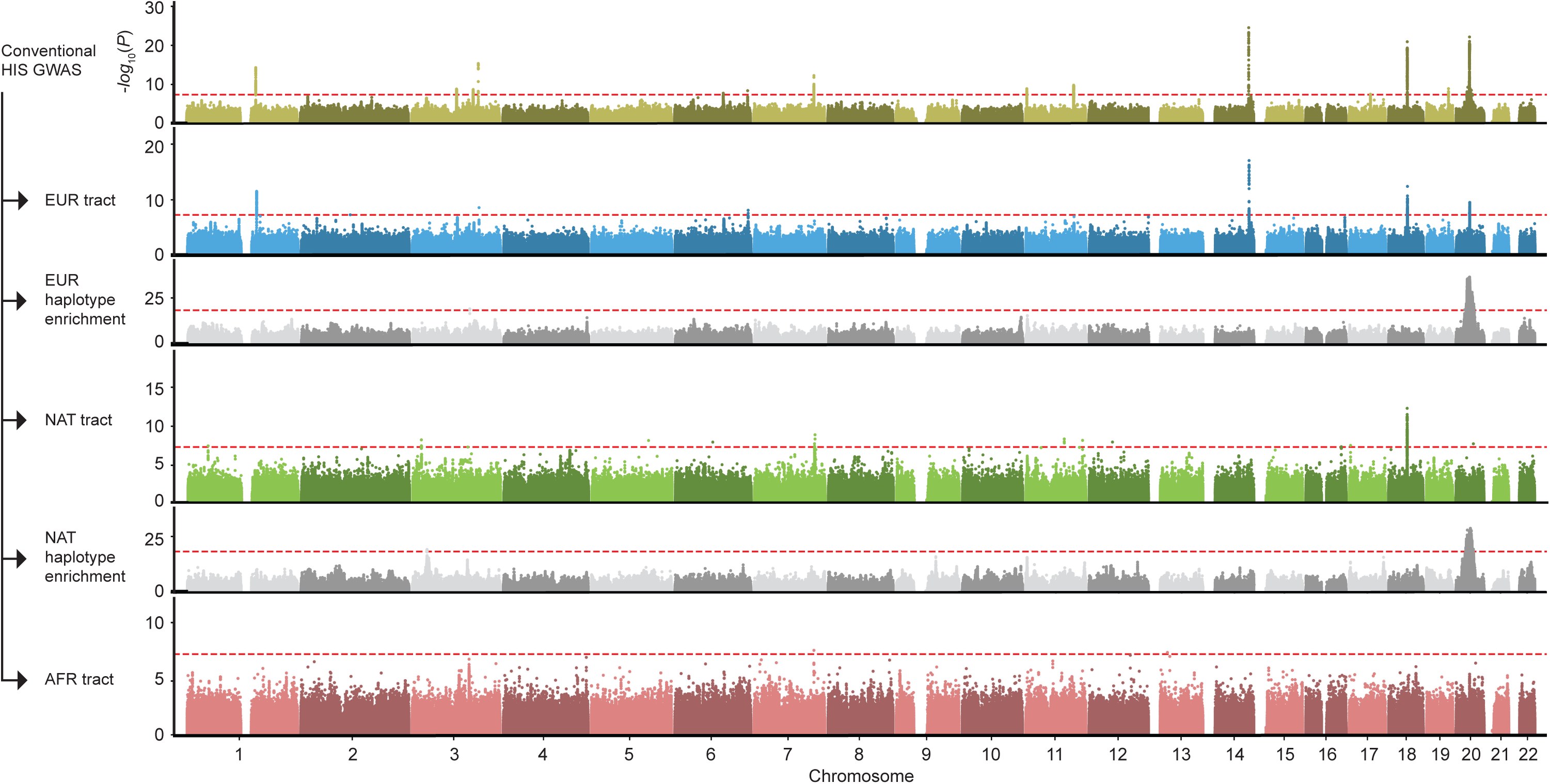
Local-ancestry-aware association analysis of Million Veteran Program (MVP) Hispanic (HIS) participants. Stacked Manhattan plots show the −log_10_(*P*) values for components of the local-ancestry-aware GWAS for MVP HIS (from top to bottom): conventional HIS GWAS, European (EUR) tract, haplotype dosage enrichment of EUR with reference to African (AFR) and Native American (NAT) haplotype dosage, NAT tract, haplotype dosage enrichment of NAT with reference to EUR and AFR haplotype dosage, and AFR tract.

**Extended Data Fig. 7.**
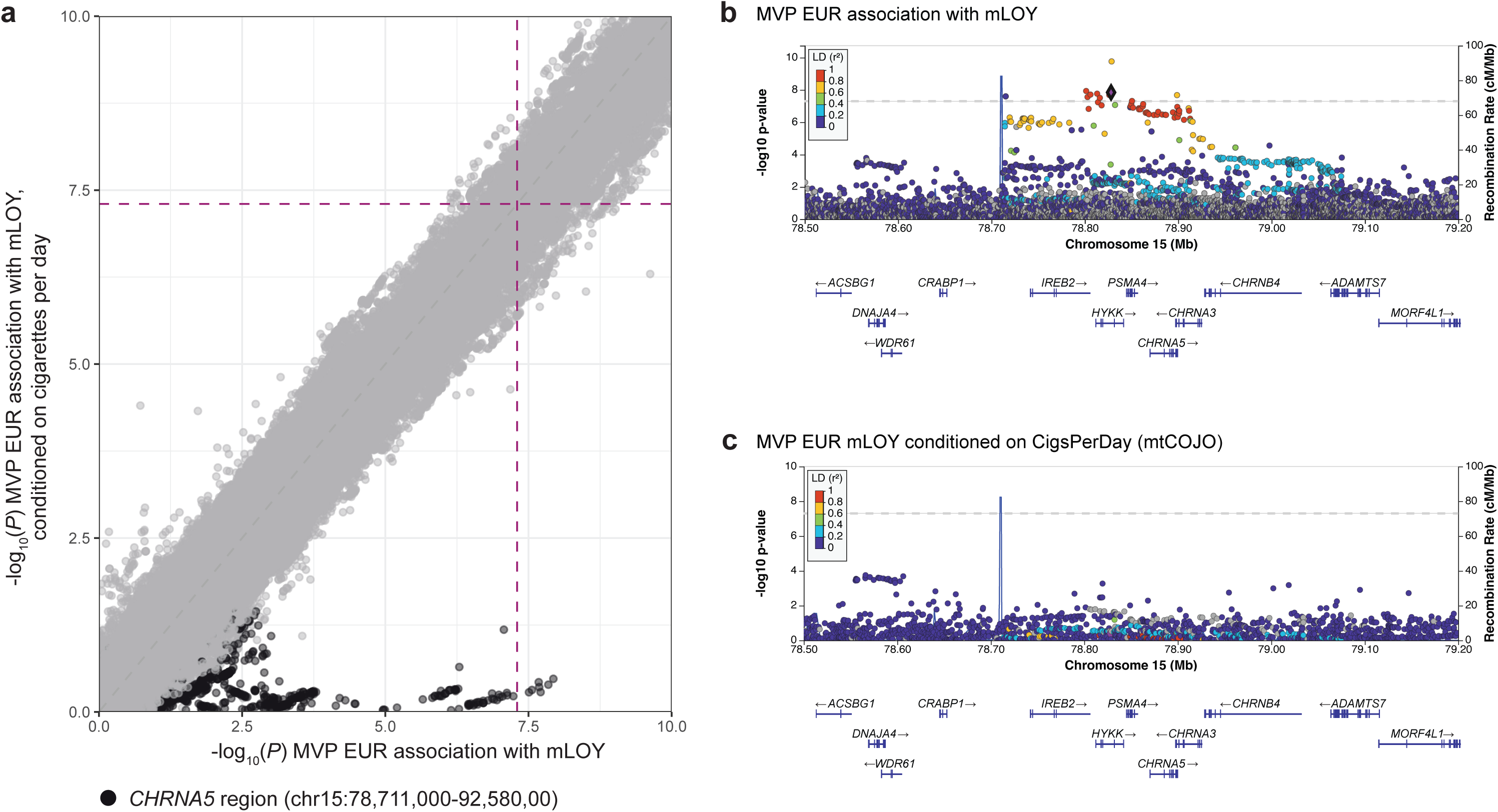
Multi-trait meta-analysis (mtCOJO) conditional analysis on smoking. **a**, Scatterplot of −log_10_(*P*) from conditioning on cigarettes-per-day with mtCOJO (LPc) vs. −log_10_(*P*) from standard mLOY GWAS (LP) in MVP European ancestry (EUR) demonstrates the region containing the *CHRNA5* gene (chr15:78,711,000-79,258,000), a known smoking locus, was primarily affected by conditioning. Red dashed lines indicate genome-wide significance at *P*<5×10^−8^. **b**, Regional Manhattan plot for *CHRNA5* in MVP EUR. **c**, Regional Manhattan plot for *CHRNA5* in MVP EUR after conditioning mLOY on cigarettes-per-day.

## Supplementary information

**Supplementary Fig. 1.**
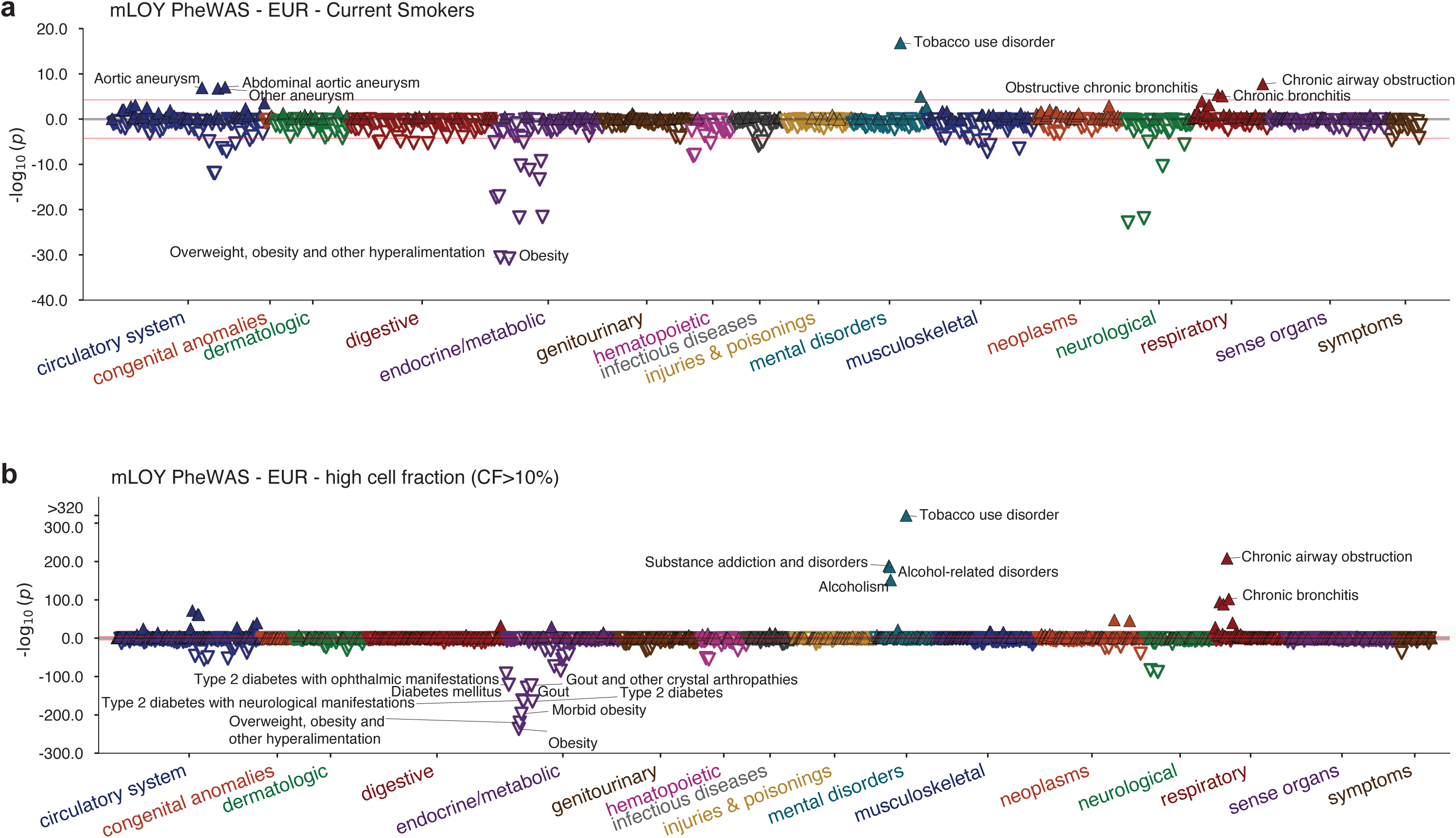
Phenome-wide association scan Manhattan plots for Million Veteran Program European ancestry. Red lines indicate significance threshold with Bonferroni correction for number of traits.

**Supplementary Fig. 2.**
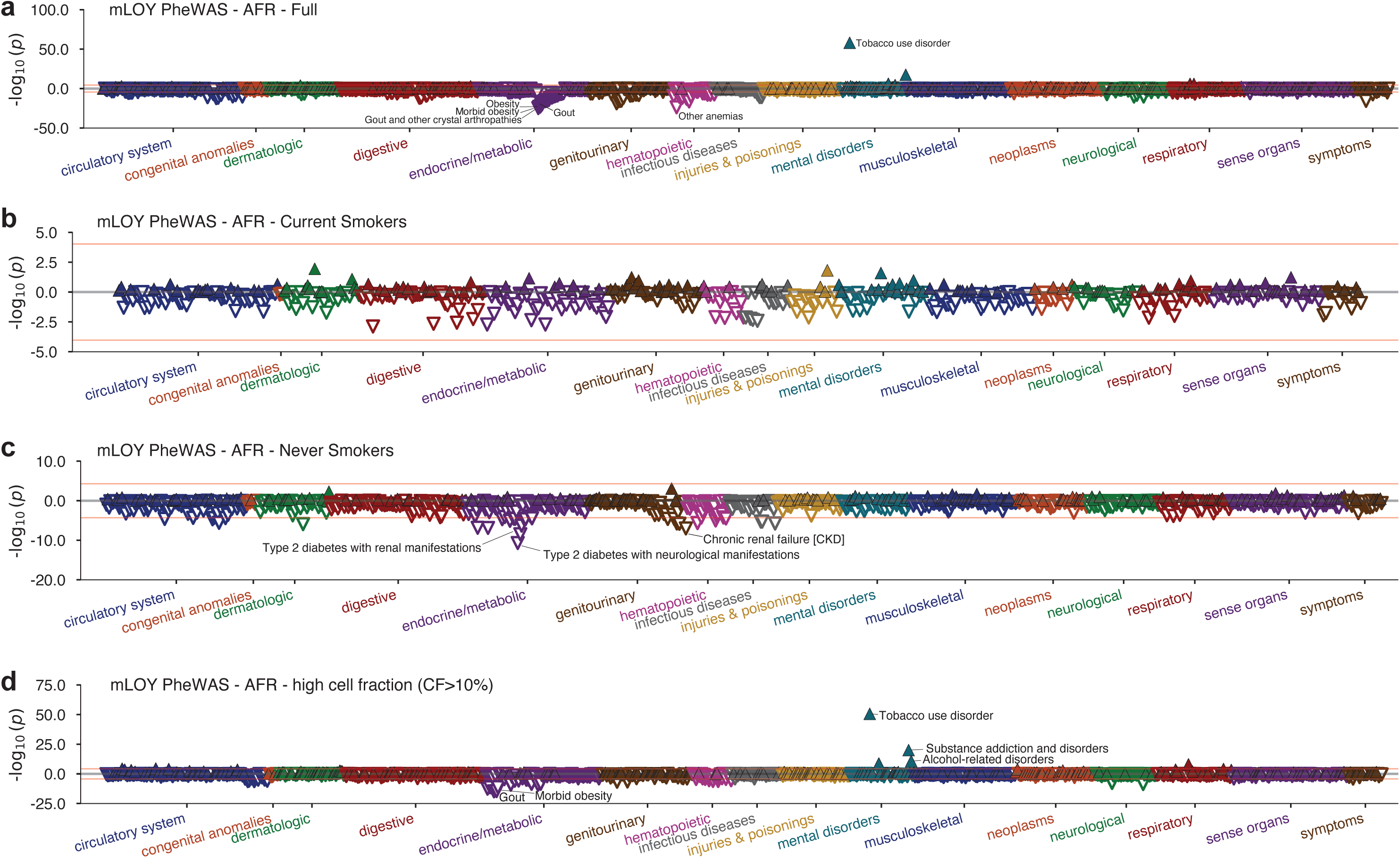
Phenome-wide association scan Manhattan plots for mosaic loss of chromosome Y in Million Veteran Program African ancestry. **a**, Full cohort; **b**, current smokers only; **c**, never smokers only; **d**, high cell fraction cases (CF>10%). Association p-values were obtained from a two-sided test of the z-statistic, calculated using logistic regression. The red line in each plot indicates Bonferroni-corrected significance.

**Supplementary Fig. 3.**
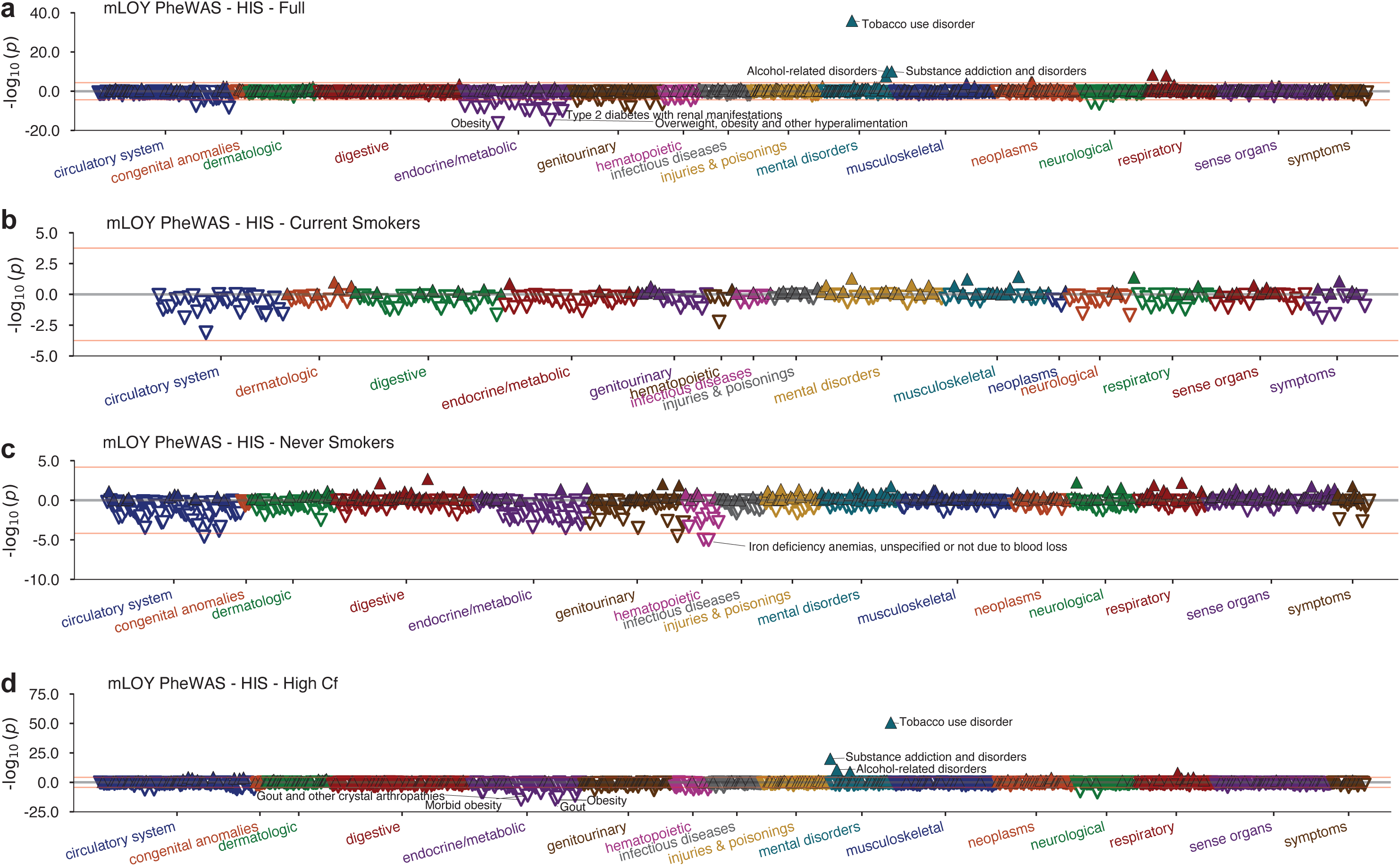
Phenome-wide association scan Manhattan plots for mosaic loss of chromosome Y in Million Veteran Program Hispanic ancestry. **a**, Full cohort; **b**, current smokers only; **c**, never smokers only; **d**, high cell fraction cases (CF>10%). Association p-values were obtained from a two-sided test of the z-statistic, calculated using logistic regression. The red line in each plot indicates Bonferroni-corrected significance.

**Supplementary Fig. 4.**
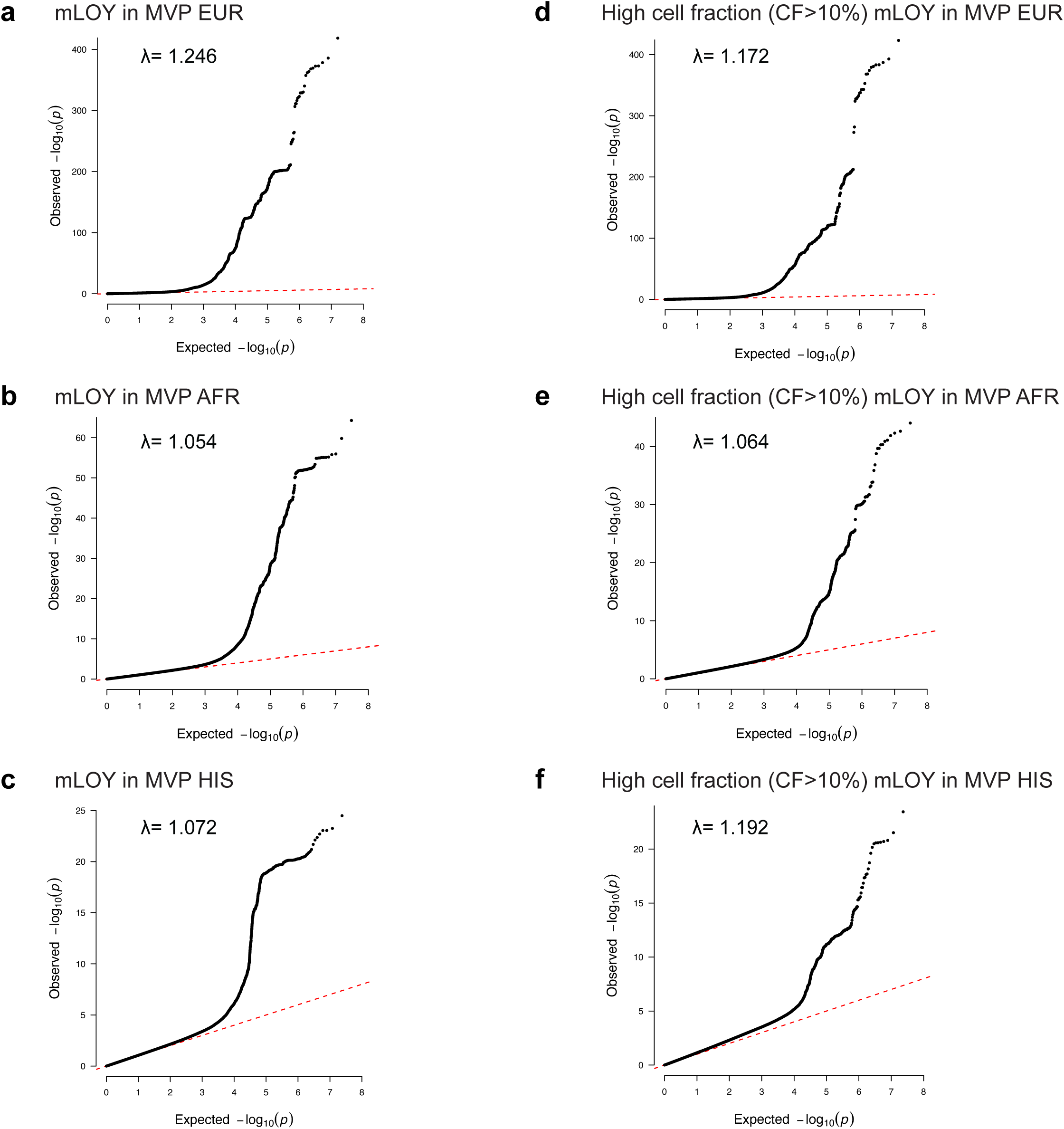
Quantile-quantile (QQ) plots of mLOY GWAS performed in this study. QQ plots show observed versus expected distributions of negative log transformed *P*-values for associations with mLOY in: **a**, Million Veteran Program (MVP) European ancestry (EUR); **b**, MVP admixed African ancestry (AFR); **c**, MVP Hispanic ancestry (HIS); **d**, High cell fraction (CF>10%) MVP EUR; **e**, High CF MVP AFR; **f**, High CF MVP HIS.

**Supplementary Fig. 5.**
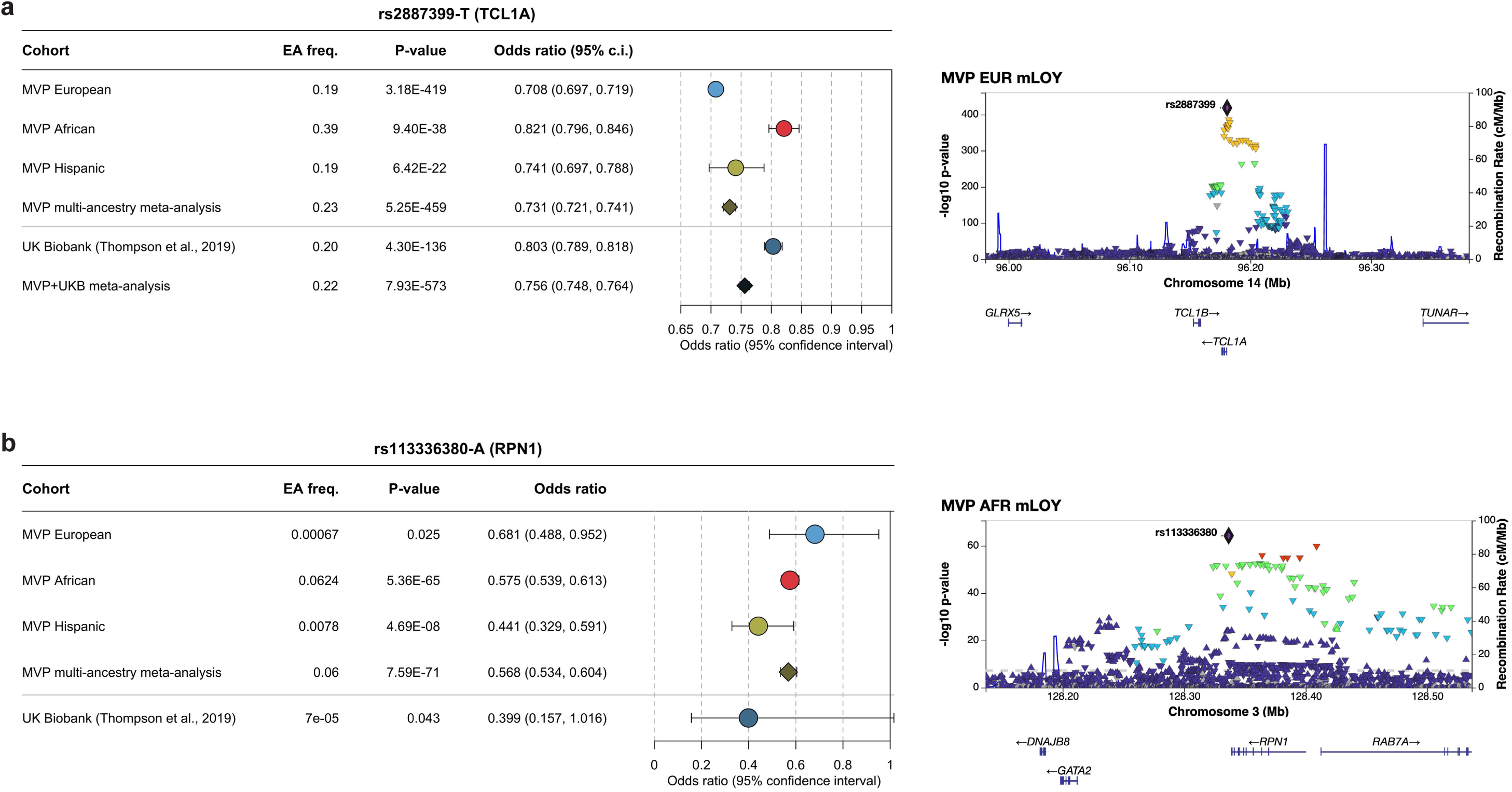
Forest plots and regional Manhattan plots for most significant mosaic loss of Y associations. **a**, rs2887399, the top SNP at *TCL1A*; **b**, rs113336380, the top African ancestry SNP at *RPN1*. Left: forest plots showing frequency of effect allele, *P*-value, and Odds ratio for each Million Veteran Program (MVP) cohort and multi-ancestry meta-analysis, plus replication cohort UK Biobank^1^ (UKB) and MVP+UKB meta-analysis. Right: Regional Manhattan plots for the MVP ancestry in which the SNP was most significant. rs113336380 was not meta-analyzed with UKB due to low frequency (MAF<0.001).

**Supplementary Fig. 6.**
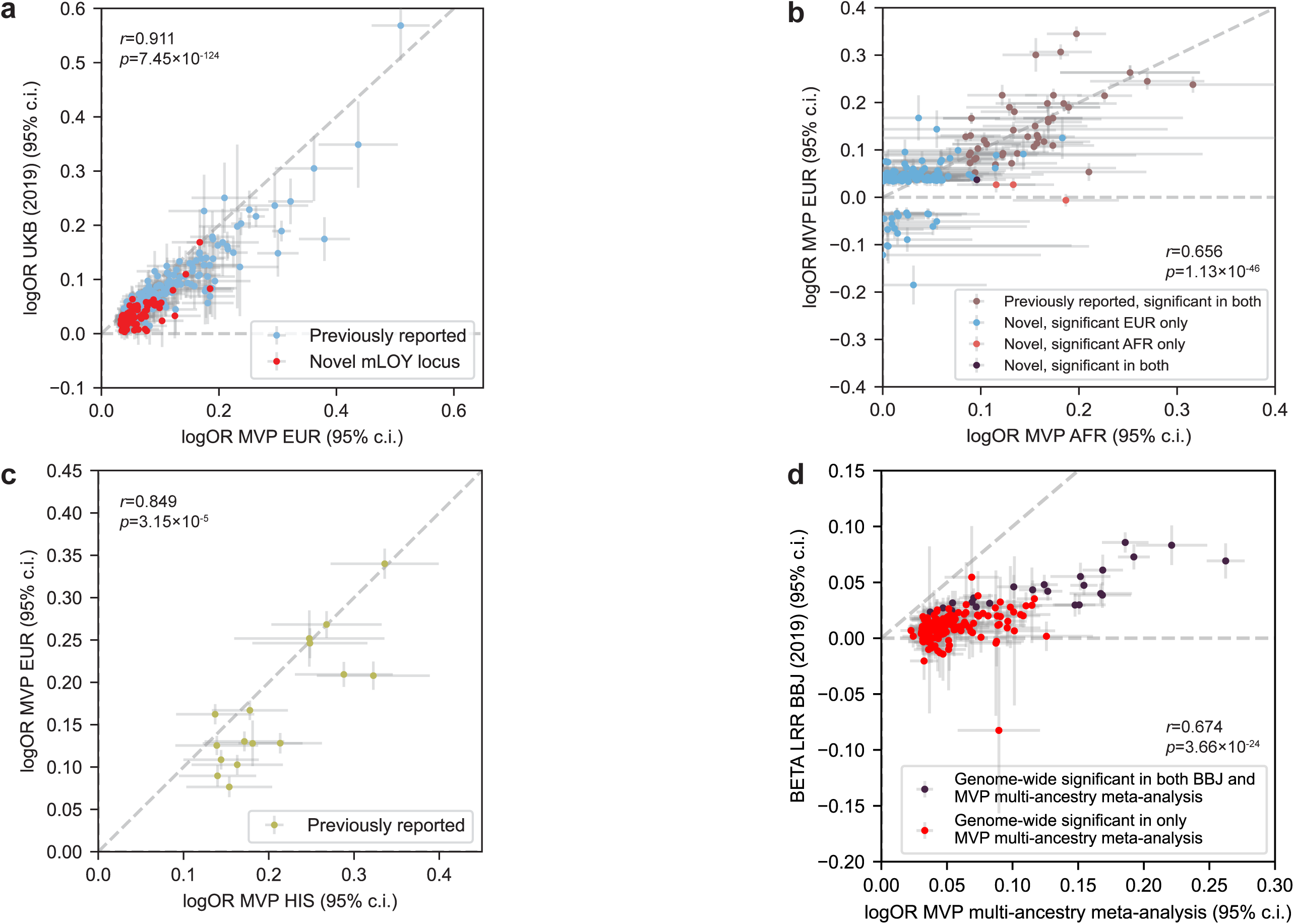
Effect comparisons between cohorts. Scatter plots comparing effect sizes (logarithm of odds ratios and 95% confidence interval) of genome-wide significant conditionally independent hits. **a**, UK Biobank^1^ versus Million Veteran Program (MVP) European (EUR), indicating previously reported loci (blue) and novel loci (pink). **b**, MVP EUR versus MVP African ancestry (AFR), indicating previously reported loci, novel loci, and novel loci exclusive to EUR or AFR. **c**, MVP EUR versus MVP Hispanic ancestry (HIS), where all HIS signals were also found in EUR. **d**, MVP multi-ancestry meta-analysis versus BioBank Japan (BBJ)^2^, indicating variants that were genome-wide significant in both studies or only MVP. BBJ effect is measured using log R ratio (LRR) intensity thresholding.

**Supplementary Figure 7.**
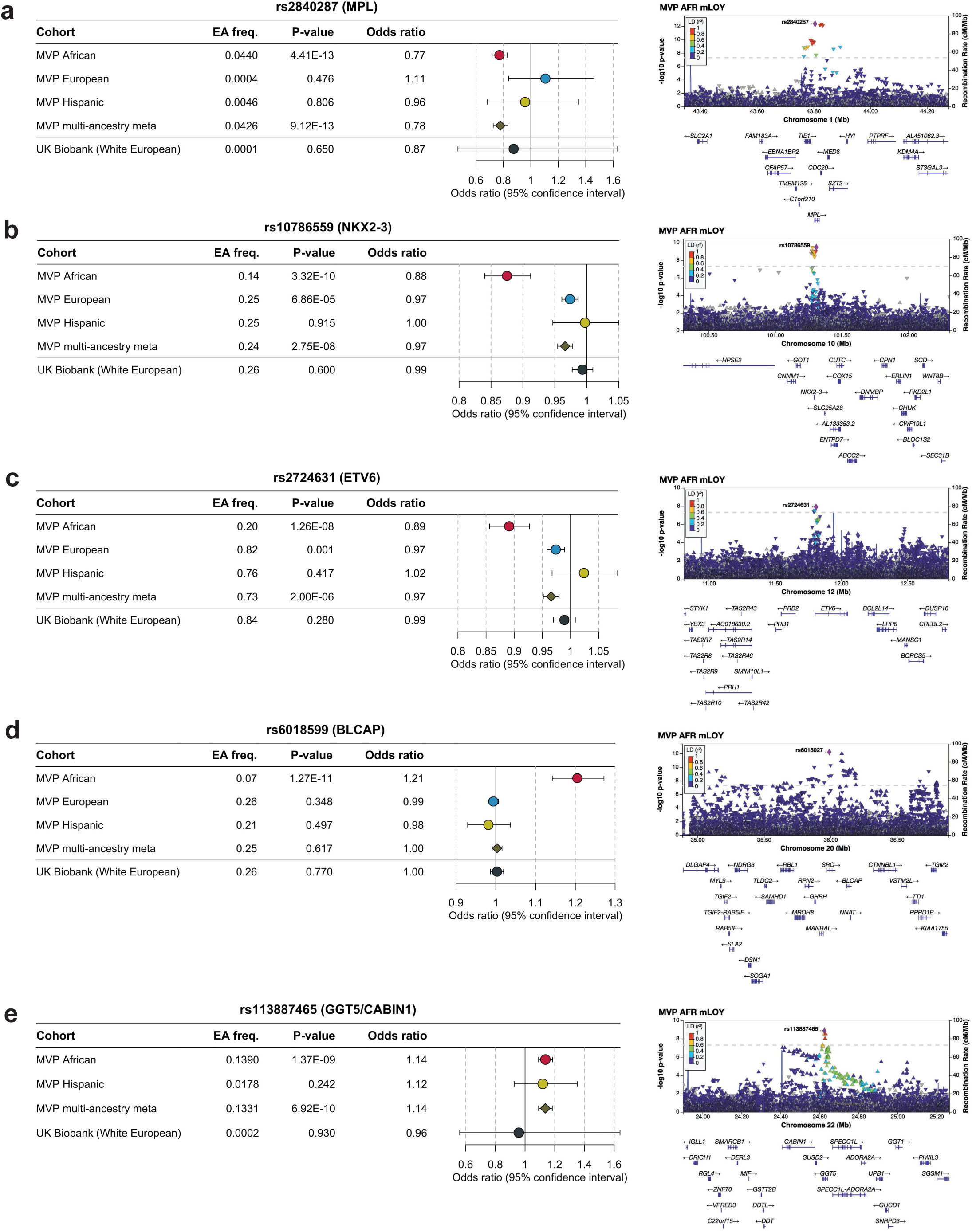
Forest plots and regional Manhattan plots for African-ancestry-specific novel mLOY loci. Five novel mLOY loci which were only genome-wide significant in Million Veteran program African Ancestry (within 1Mb): **a,** *MPL*, **b,** *NKX2-3*, **c,** *ETV6*, **d,** *BLCAP*, and **e,** *GGT5*/*CABIN1* (not available in MVP European).

**Supplementary Fig. 8.**
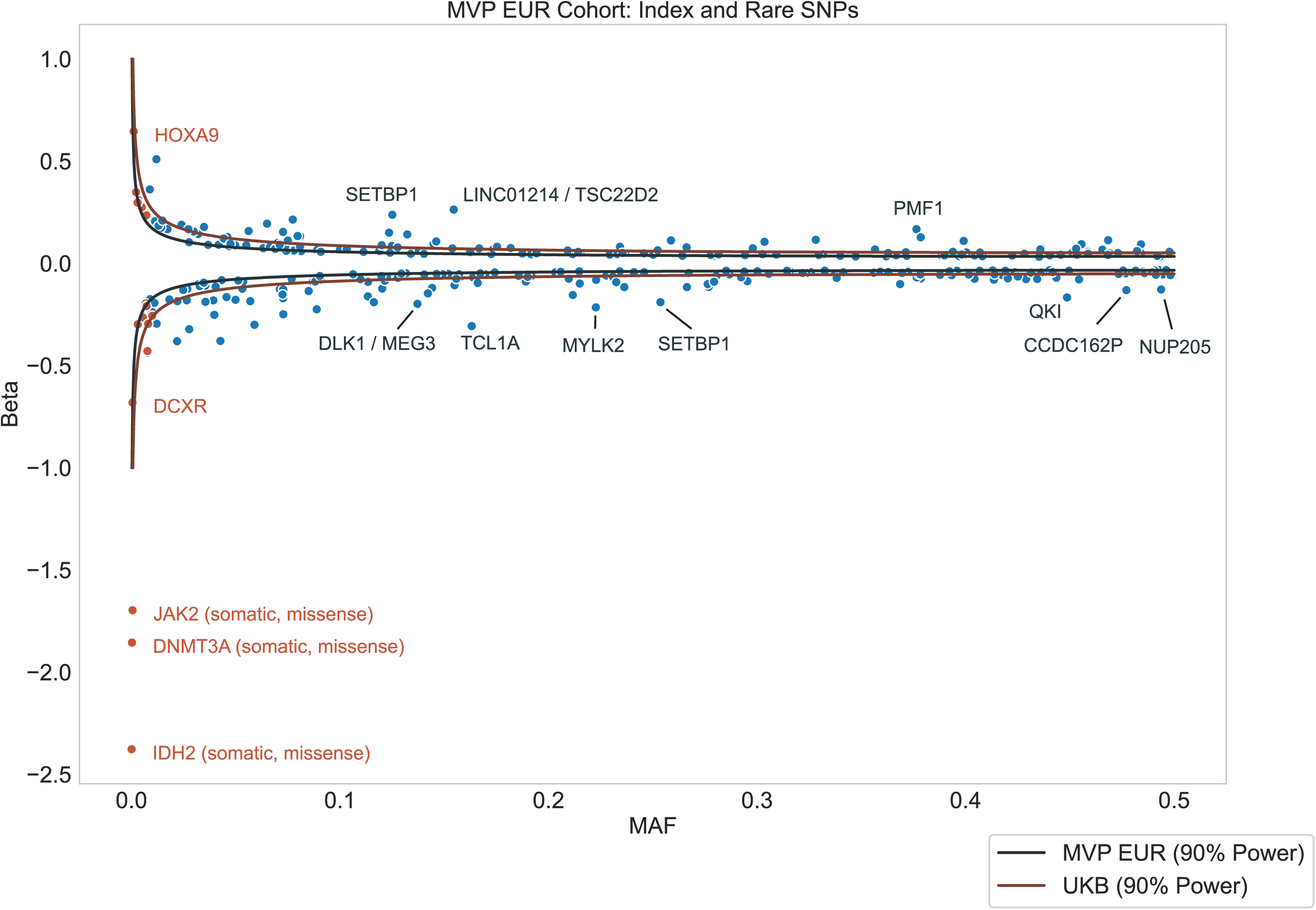
Relationship between minor allele frequency and effect size. For 327 genome-wide significant conditionally independent SNPs, plotting effect size (beta in log odds) versus minor allele frequency (MAF). Simulated power curves are shown for MVP and UKB^1^ MAFs representing the expected magnitude of beta detectable at *P*<5×10^−8^. Significant rare variants (MAF<0.001 in MVP) are shown in orange. Gene names provided for SNPs with *P*<1×10^−100^.

**Supplementary Fig. 9.**
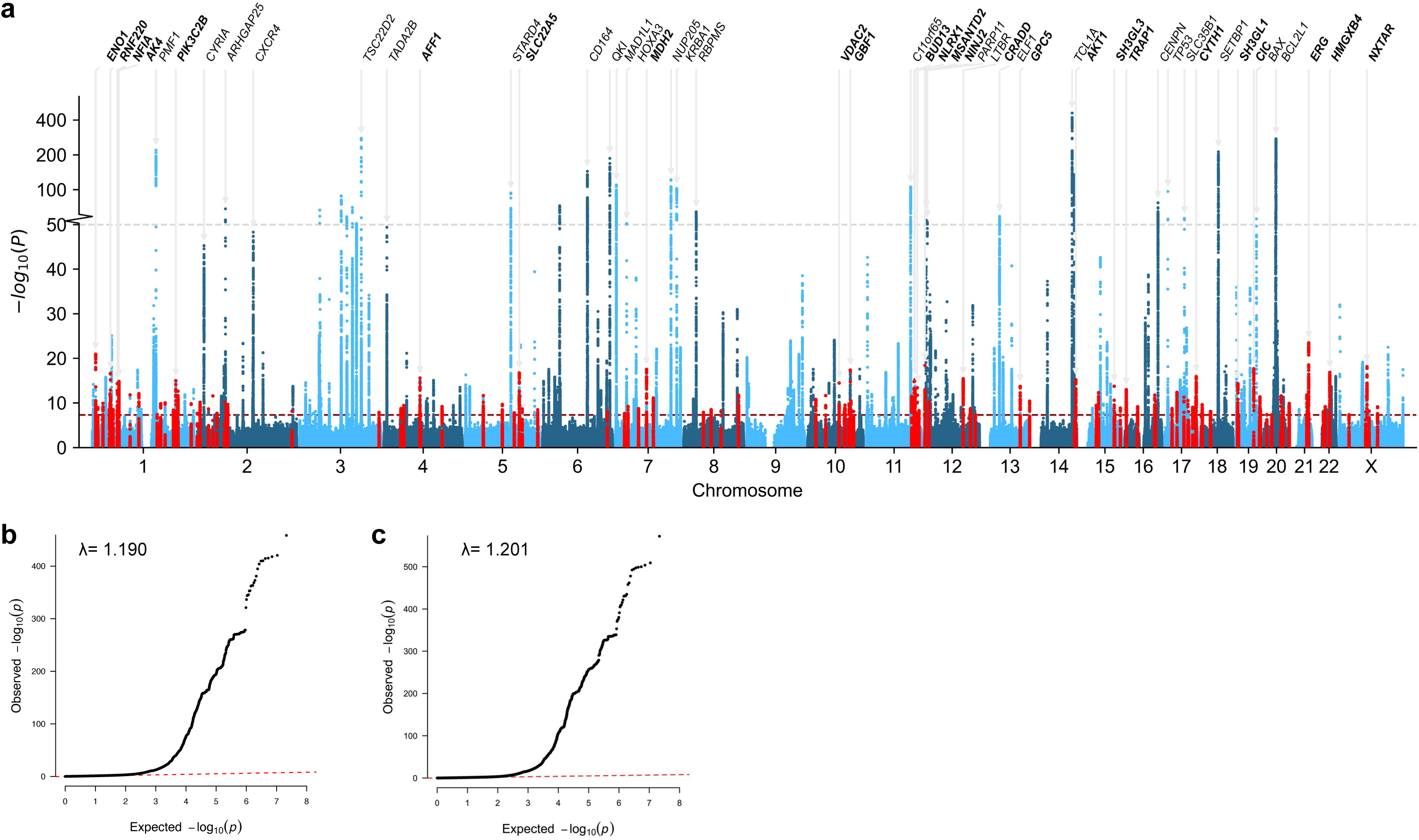
Manhattan and quantile-quantile (QQ) plots of Million Veteran Program (MVP) multi-ancestry meta-analysis. **a**, Manhattan plot shows the −log10(*P*) for associations of genetic variants with mLOY in the MVP meta-analysis of European, African, and Hispanic ancestries. Novel mLOY index variants and variants within ± 50 Kb are highlighted in red. The red line indicates the genome-wide significance threshold (*P*<5×10^−8^). The grey dotted line represents a transition from linear to log-scale on the y-axis.The 25 most significant known and novel loci are annotated with the nearest gene. **b**, QQ plot shows observed versus expected distributions of *P*-values in MVP multi-ancestry meta-analysis. **c**, QQ plot shows observed versus expected distributions of *P*-values in MVP and UK Biobank meta-analysis (corresponding to the Manhattan plot shown in Main Fig. 2d).

**Supplementary Fig. 10.**
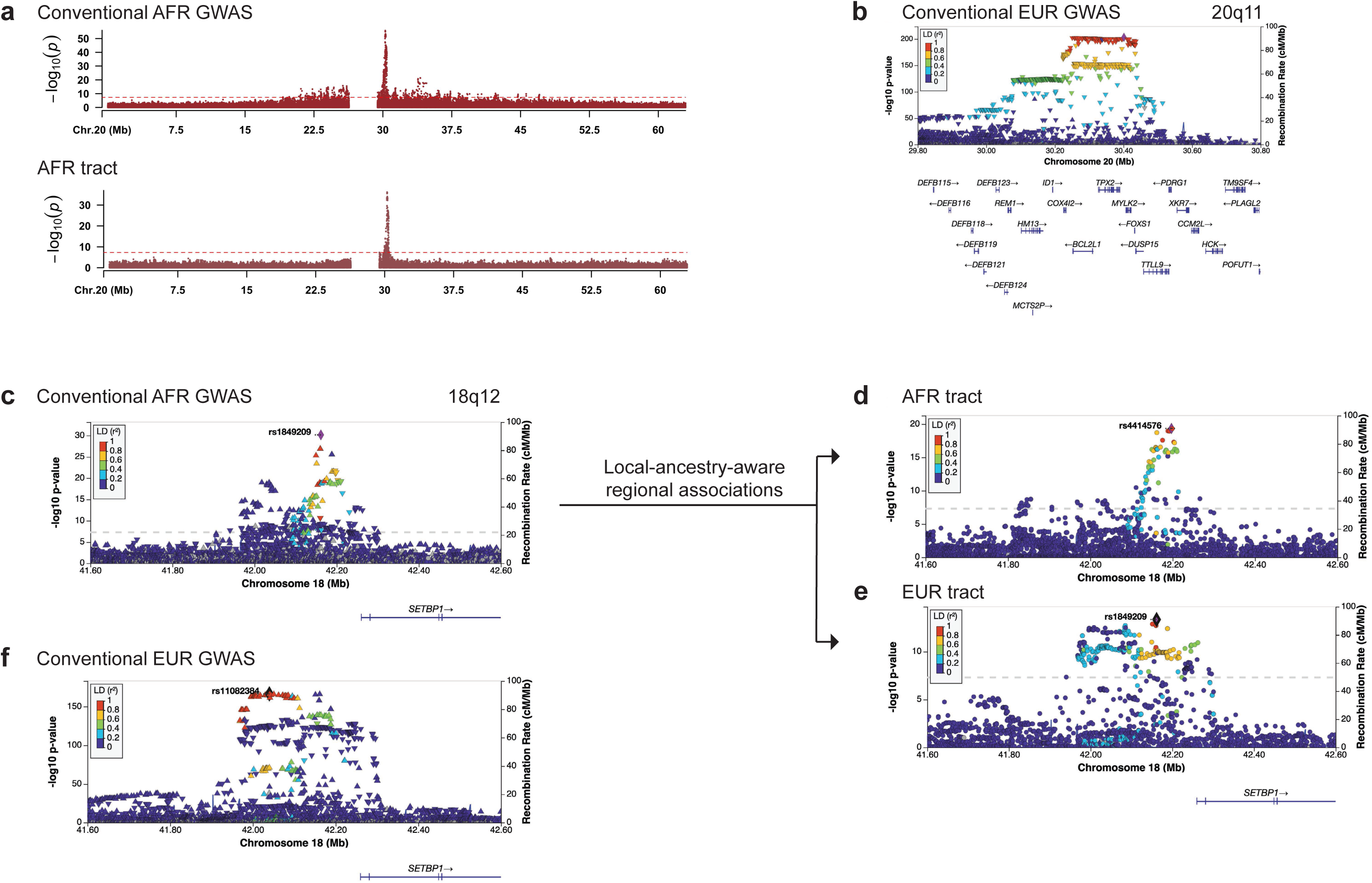
Supporting evidence in Million Veteran Program admixed African samples for local ancestry associations with mLOY. a, Chromosome 20 Manhattan plots for conventional admixed African (AFR) and AFR tract association *P*-values. **b,** Conventional European (EUR) regional Manhattan plot for the *BCL2L1* locus. **c,** At 18q12 (*SETBP1*), we observed complex linkage disequilibrium structure; this was resolved by deconvolving the **d,** AFR and **e,** EUR tracts. **f,** Regional Manhattan plot for *SETBP1* in conventional EUR association analysis.

**Supplementary Fig. 11.**
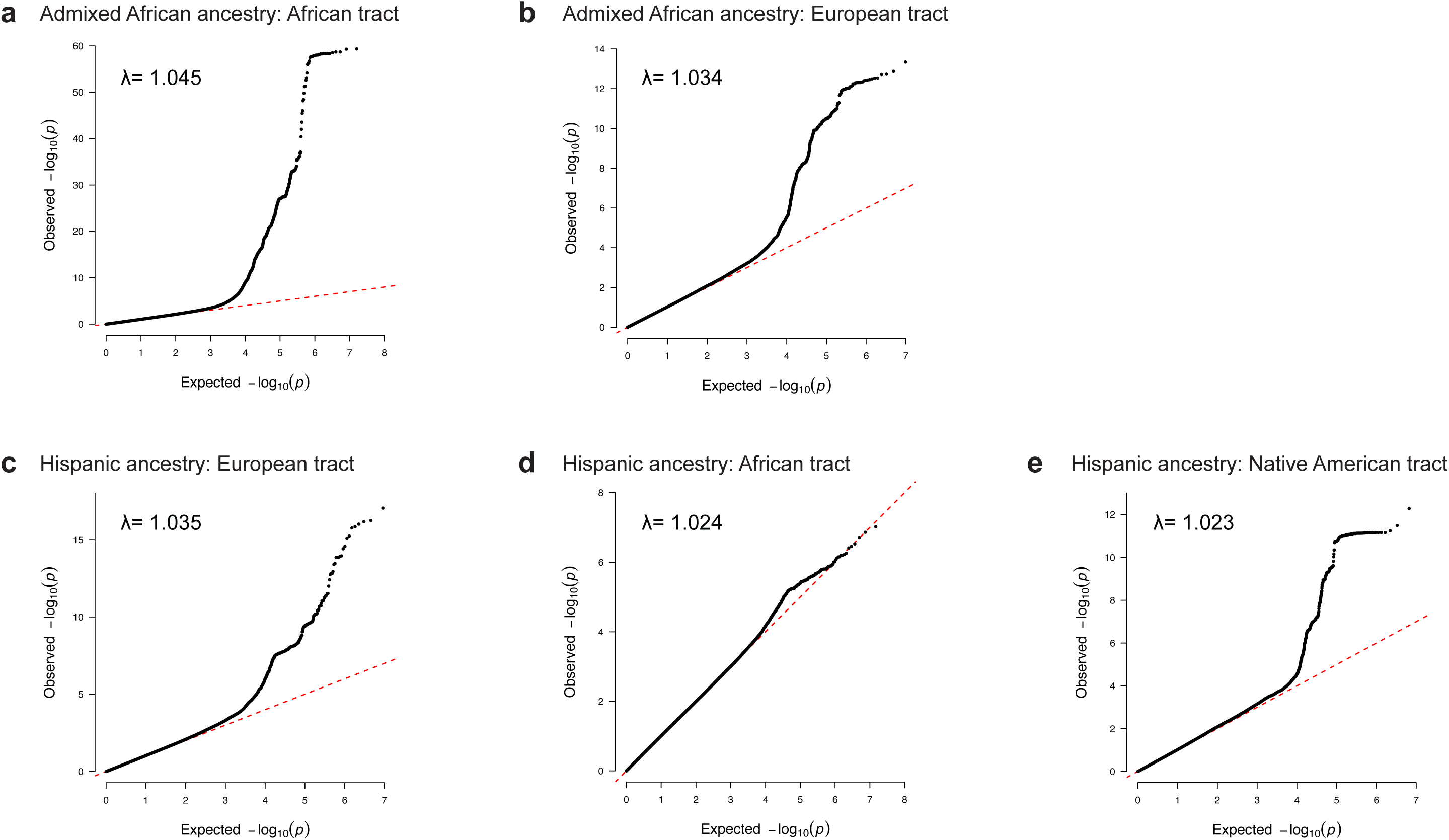
Local ancestry tract quantile-quantile (QQ) plots. QQ plots show observed versus expected distributions of *P*-values in Million Veteran Program (MVP) admixed African tracts for **a**, African, and **b**, European ancestral contributions; and in MVP Hispanic tracts for **c**, European, **d**, African, and **e**, Native American ancestral contributions.

**Supplementary Fig. 12.**
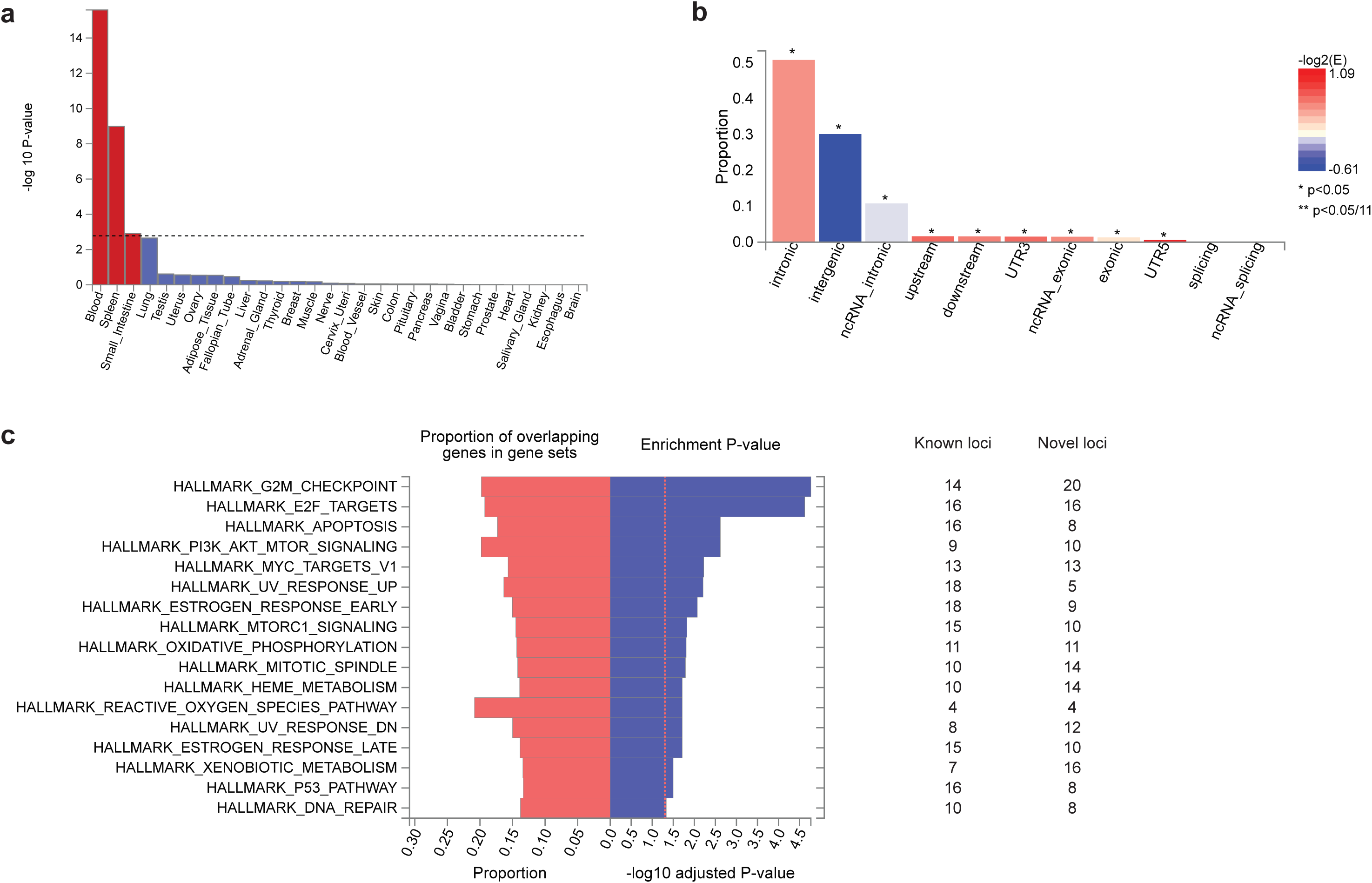
Gene set enrichment of mLOY associations. For genes positionally mapped to significant mLOY loci in the Million Veteran Program + UK Biobank meta-analysis: **a,** −log_10_(*P*) values for enrichment of GTEx v8 tissue types. **b**, Functional consequences of SNPs on genes, and **c,** FDR adjusted −log_10_(*P*) for enrichment of Hallmark gene sets, showing counts of known and novel loci in each gene set.

**Supplementary Fig. 13.**
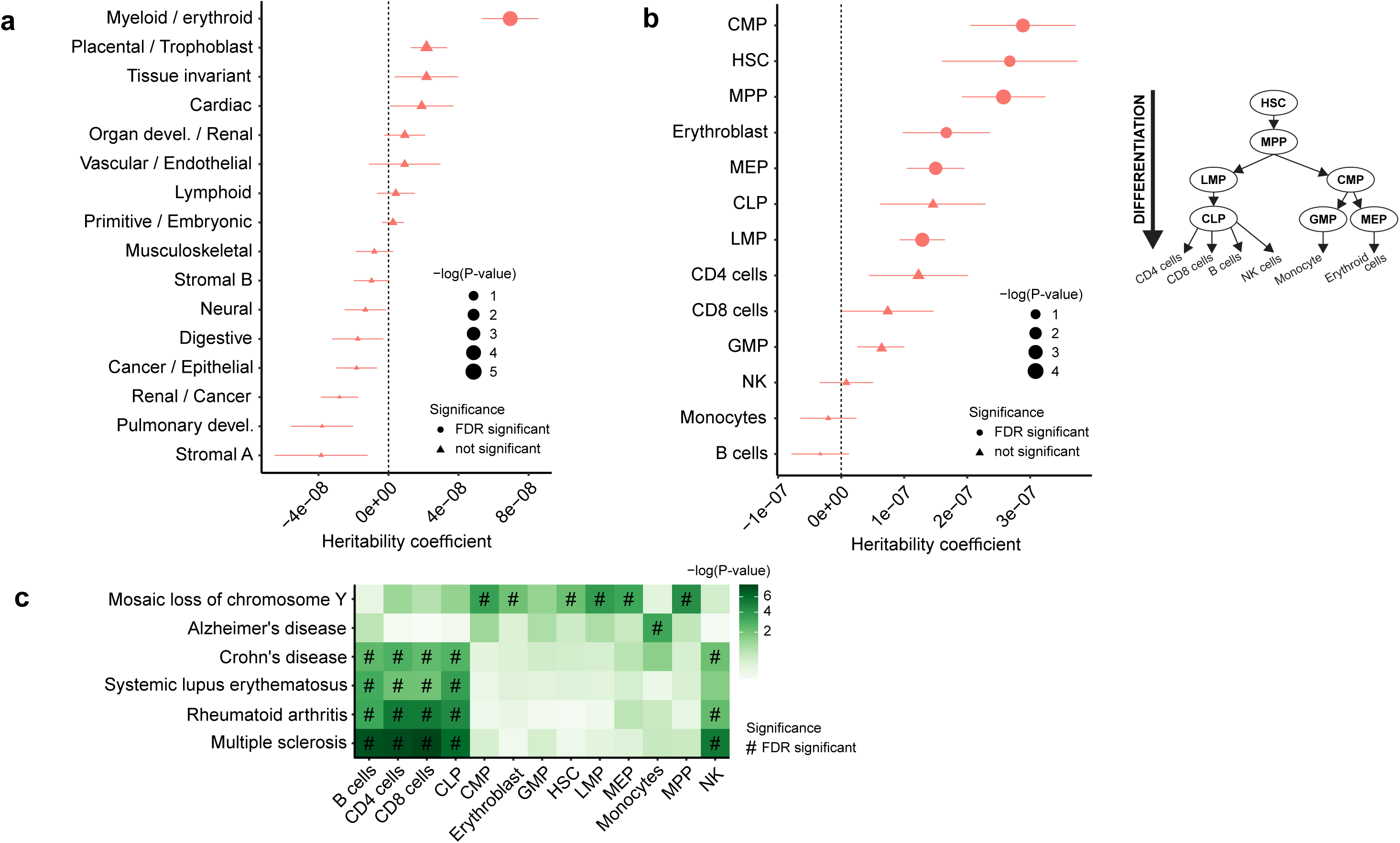
Genomic enrichment of mLOY risk variants in open chromatin regions across human cell types. (a-. **b)** Enrichment of mLOY risk variants in cell-specific open chromatin regions from **a,** main human body lineages and **b,** blood lineages. Symbols (circle/triangle) represent the LD score coefficient and horizontal bars reflect standard error. The positive LD score coefficient signifies enrichment in heritability (horizontal bars reflect standard error). Symbol size reflects the value of LD score regression. (Right) Diagram illustrating the hierarchical structure of the human hematopoietic system, highlighting the blood lineages from the atlas. **c,** Comparison of enrichment of blood lineages across a panel of diseases. CLP, common lymphoid progenitor cells; CMP, common myeloid progenitor cells; GMP, granulocyte-macrophage progenitor cells; HSC, hematopoietic stem cells; LMP, lymphoid-primed progenitor cells; MEP, megakaryote-erythroid progenitor cells; MMP, multipotent progenitor cells; NK, natural killer cells.

**Supplementary Fig. 14.**
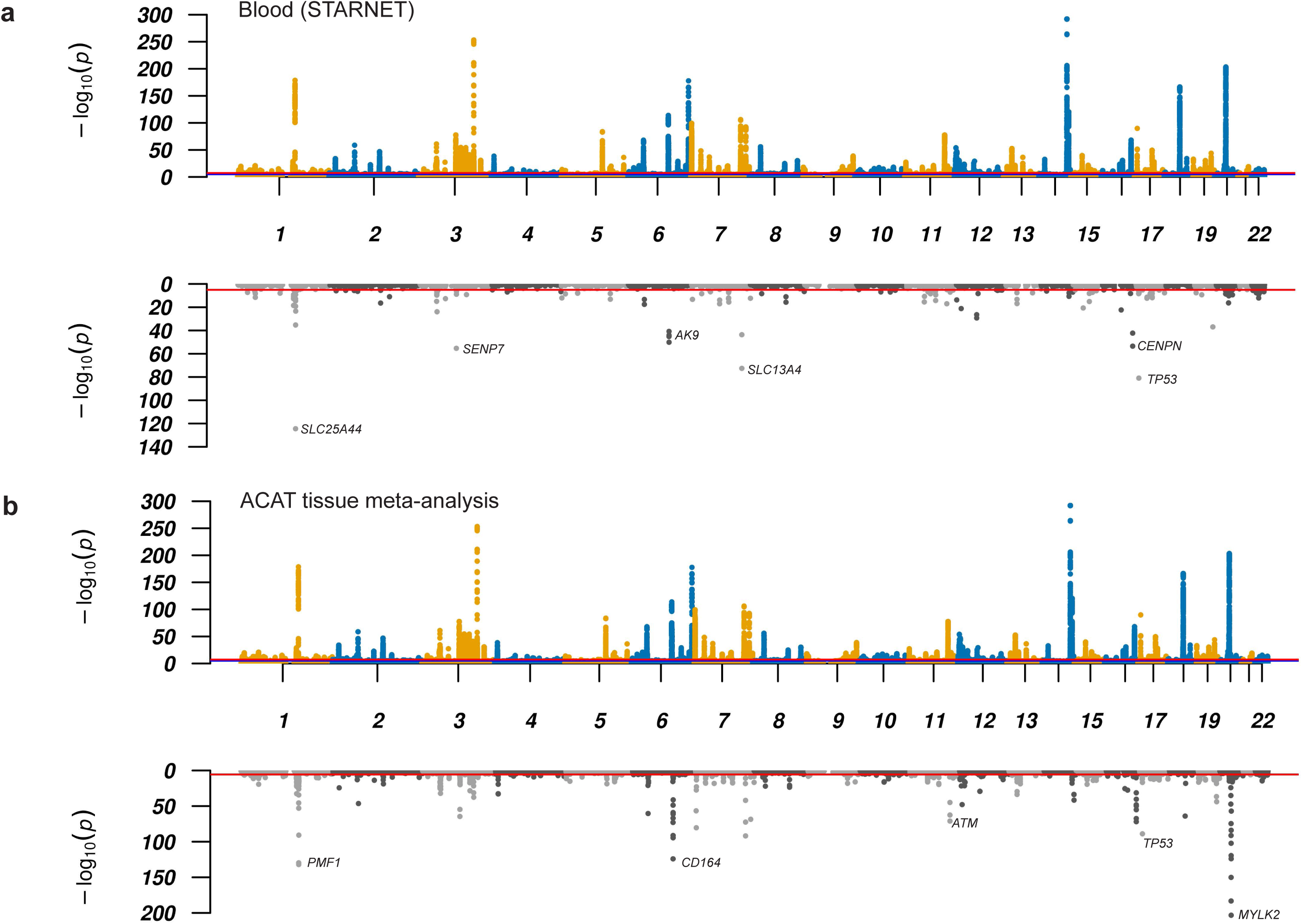
Global Miami plots for transcriptome-wide association study. Miami plots with Million Veteran Program European participants −log10(*P*) for associations of genetic variants from GWAS (top) and −log10(*P*) for associations of gene features from TWAS (bottom). Plots show TWAS results for **a**, Blood (STARNET) and **b**, ACAT tissue meta-analysis.

**Supplementary Figure 15.**
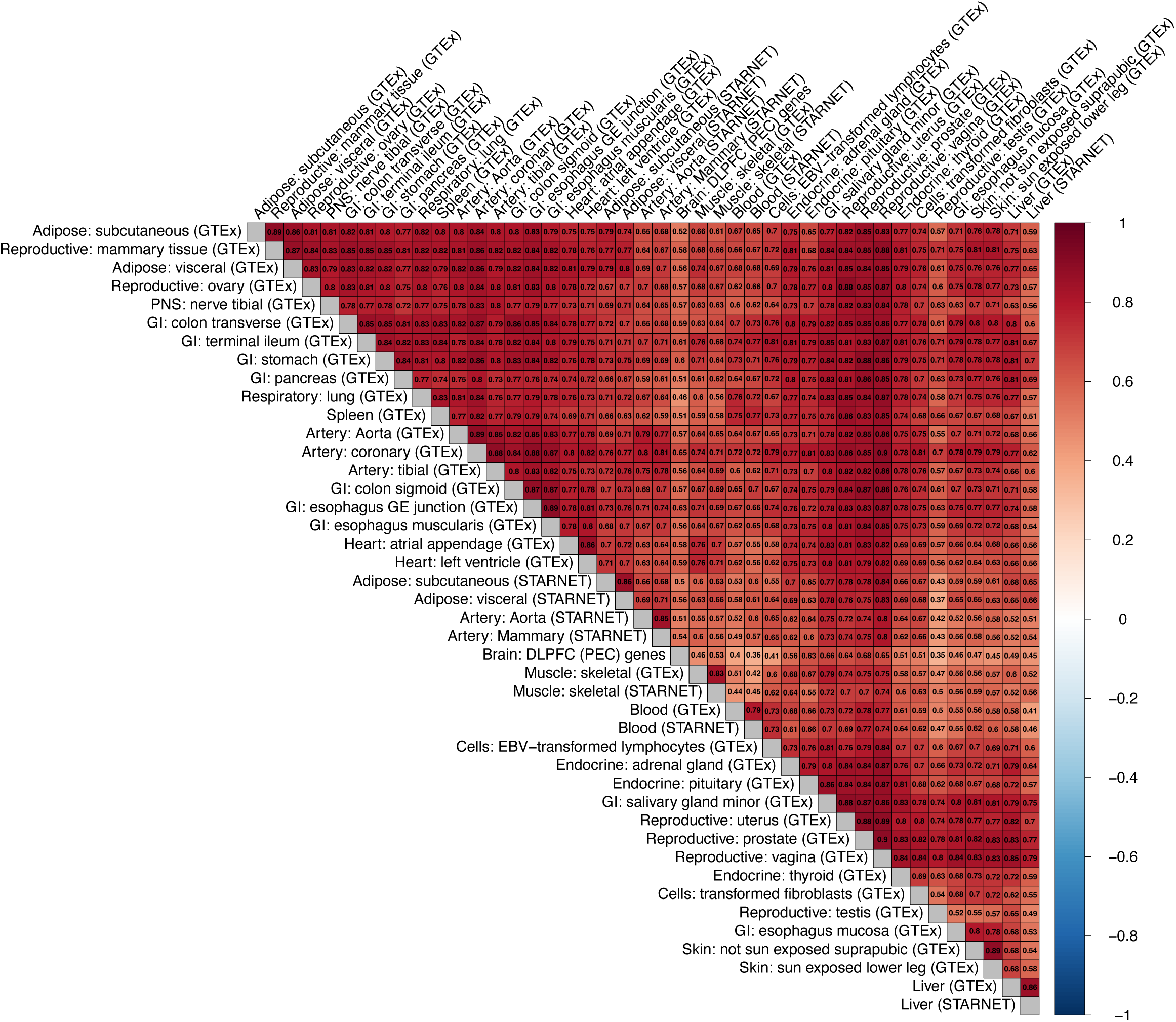
Correlation of gene expression across tissues from transcriptome-wide association study (TWAS) Values represent Pearson correlation coefficient for imputed expression changes.

**Supplementary Fig. 16.**
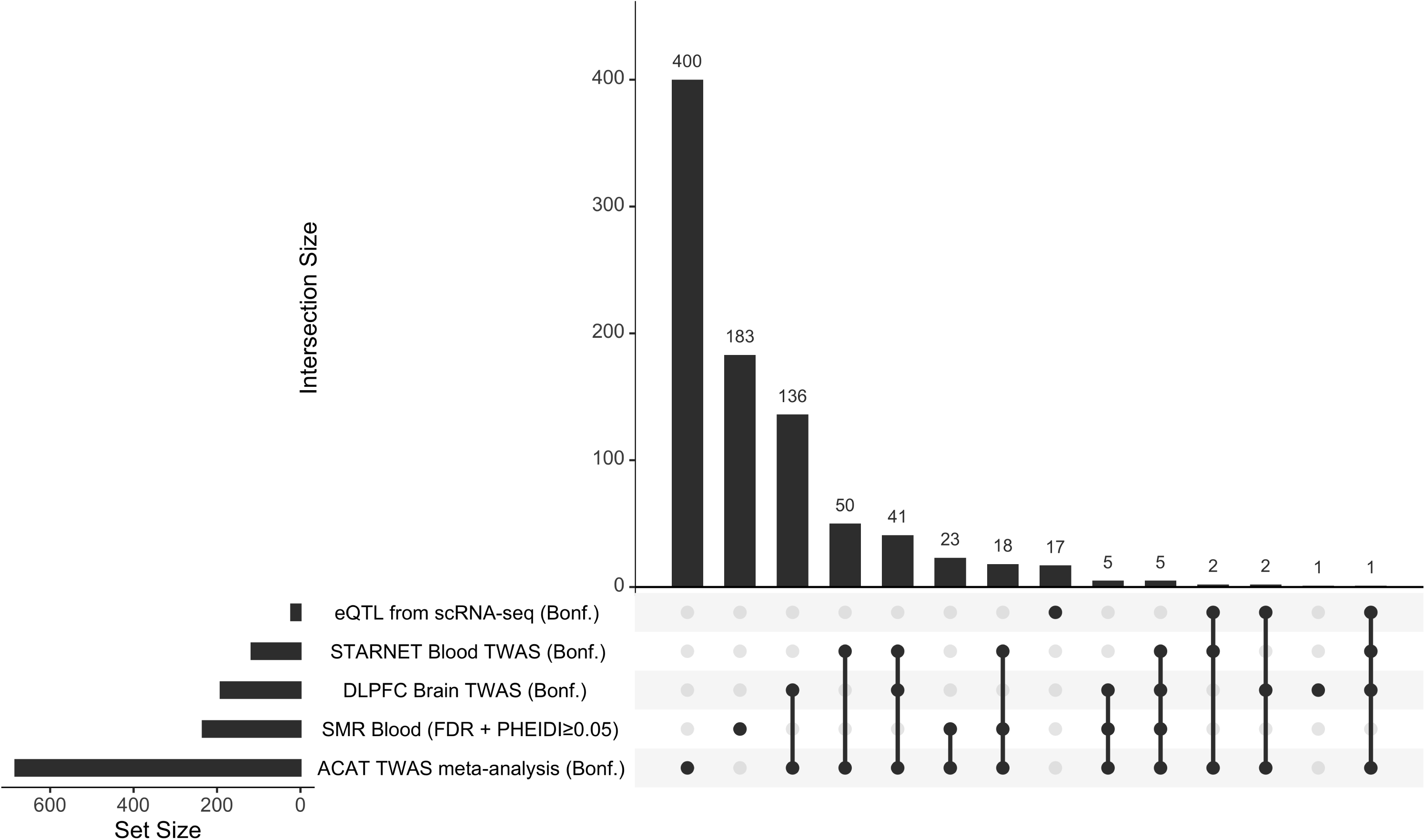
UpSet plot showing overlap between gene transcript association methods. Genes are derived from eQTLs with single cell RNA-seq (scRNA-seq) data across different immune cell populations, STARNET Blood transcriptome-wide association study (TWAS), dorsolateral prefrontal cortex TWAS, Summary Mendelian Randomization (SMR), and ACAT TWAS meta-analysis. Bonf., Bonferroni-corrected significance; FDR, false discovery rate.

**Supplementary Fig. 17.**
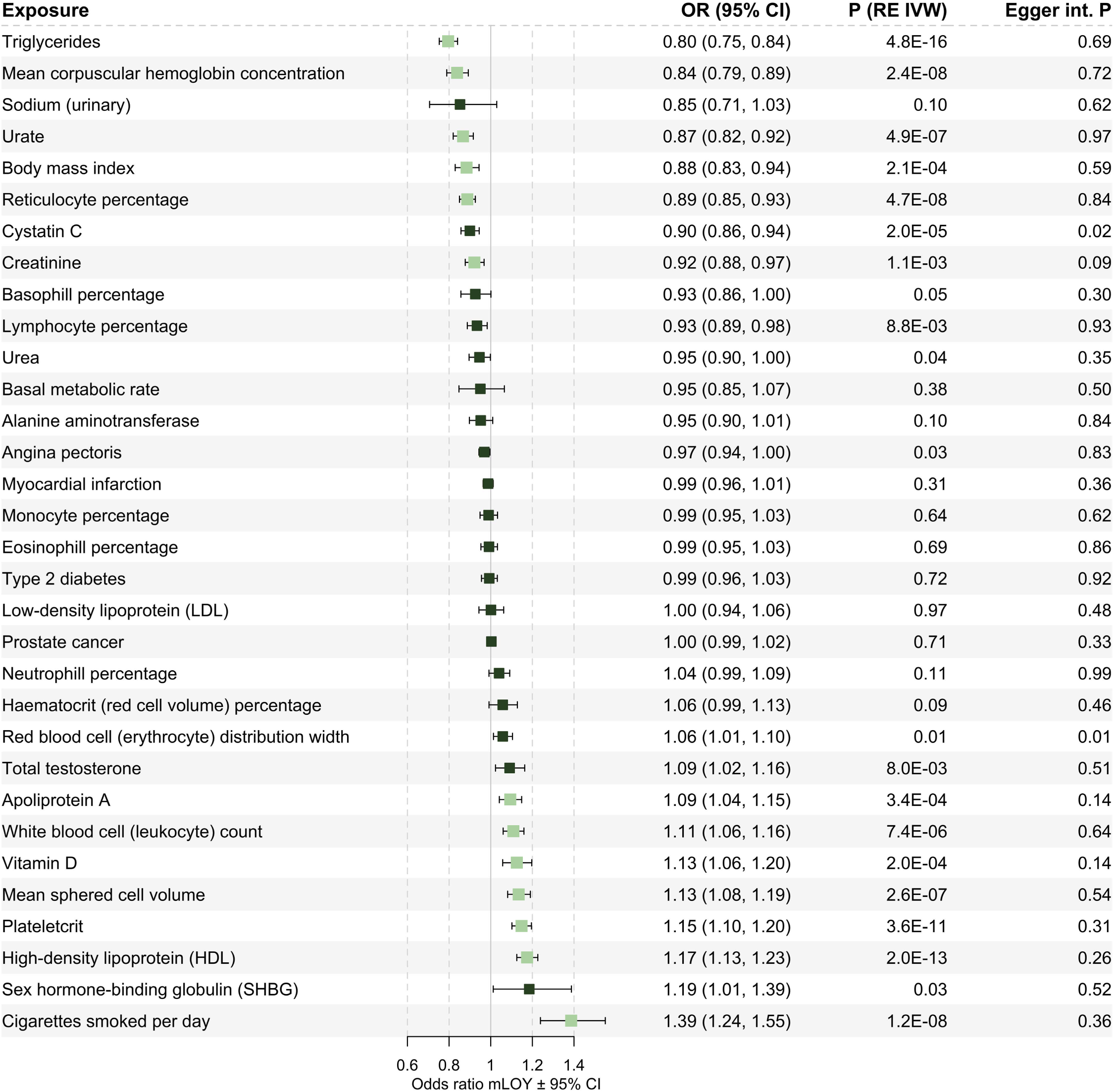
Forward Mendelian Randomization (MR) for exposure traits on mLOY. Forest plots showing random effects inverse-variance weighted (RE IVW) MR for 32 exposure traits on mLOY outcome in Million Veteran Program Europeans. Lighter box color denotes the exposure reached Bonferroni-corrected significance in RE IVW (*P*<0.05/32) and *P* MR-Egger intercept (int.)≥0.05. OR, odds ratio; CI, confidence interval.

**Supplementary Fig. 18.**
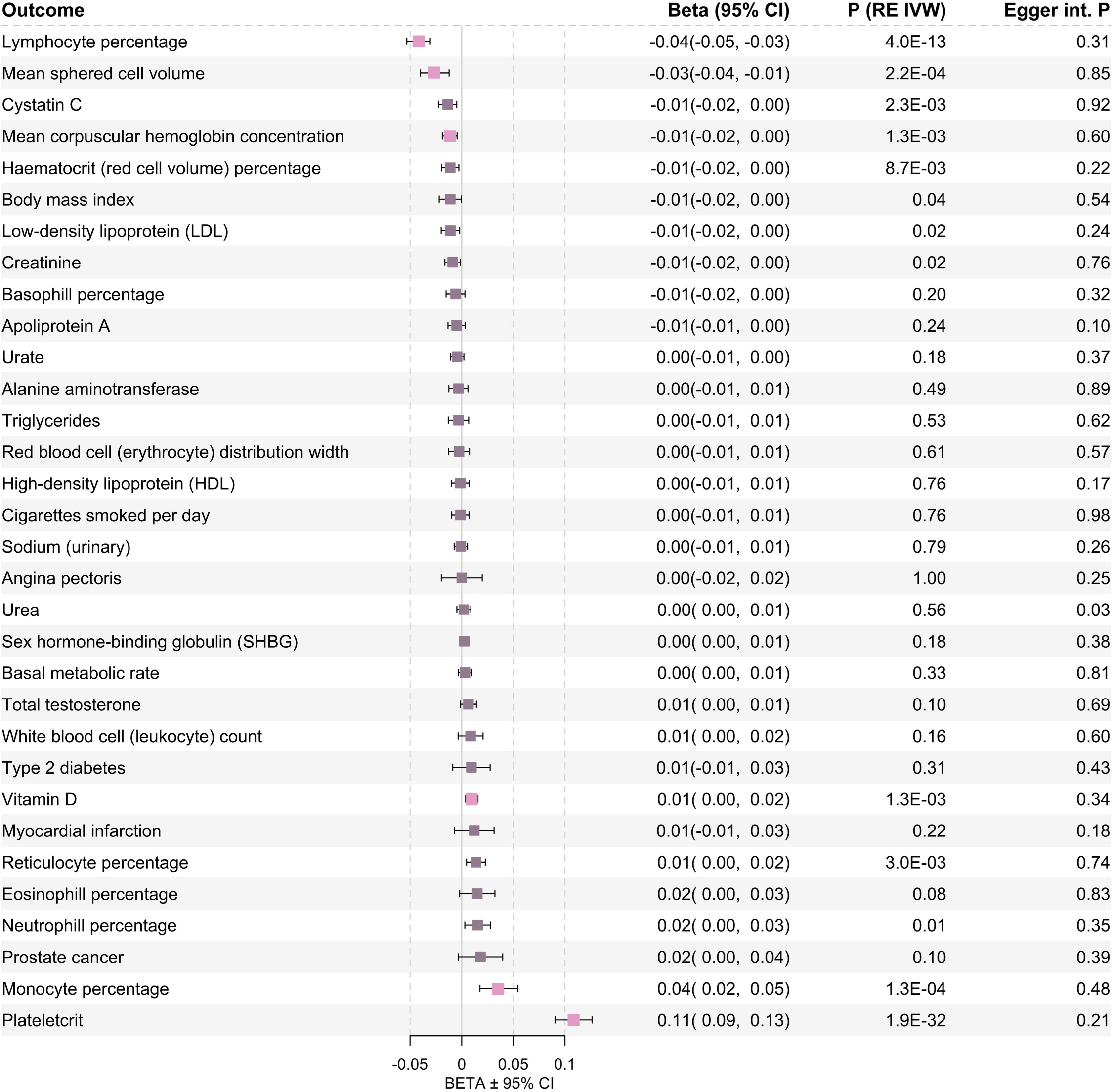
Reverse Mendelian Randomization (MR) for mLOY exposure on all trait outcomes. Forest plots showing random effects inverse-variance weighted (RE IVW) MR for mLOY exposure on 32 trait outcomes in Million Veteran Program Europeans. Lighter box color denotes the exposure reached Bonferroni-corrected significance in RE IVW (*P*<0.05/32) and *P* MR-Egger intercept (int.)≥0.05. OR, odds ratio; CI, confidence interval.

**Supplementary Fig. 19.**
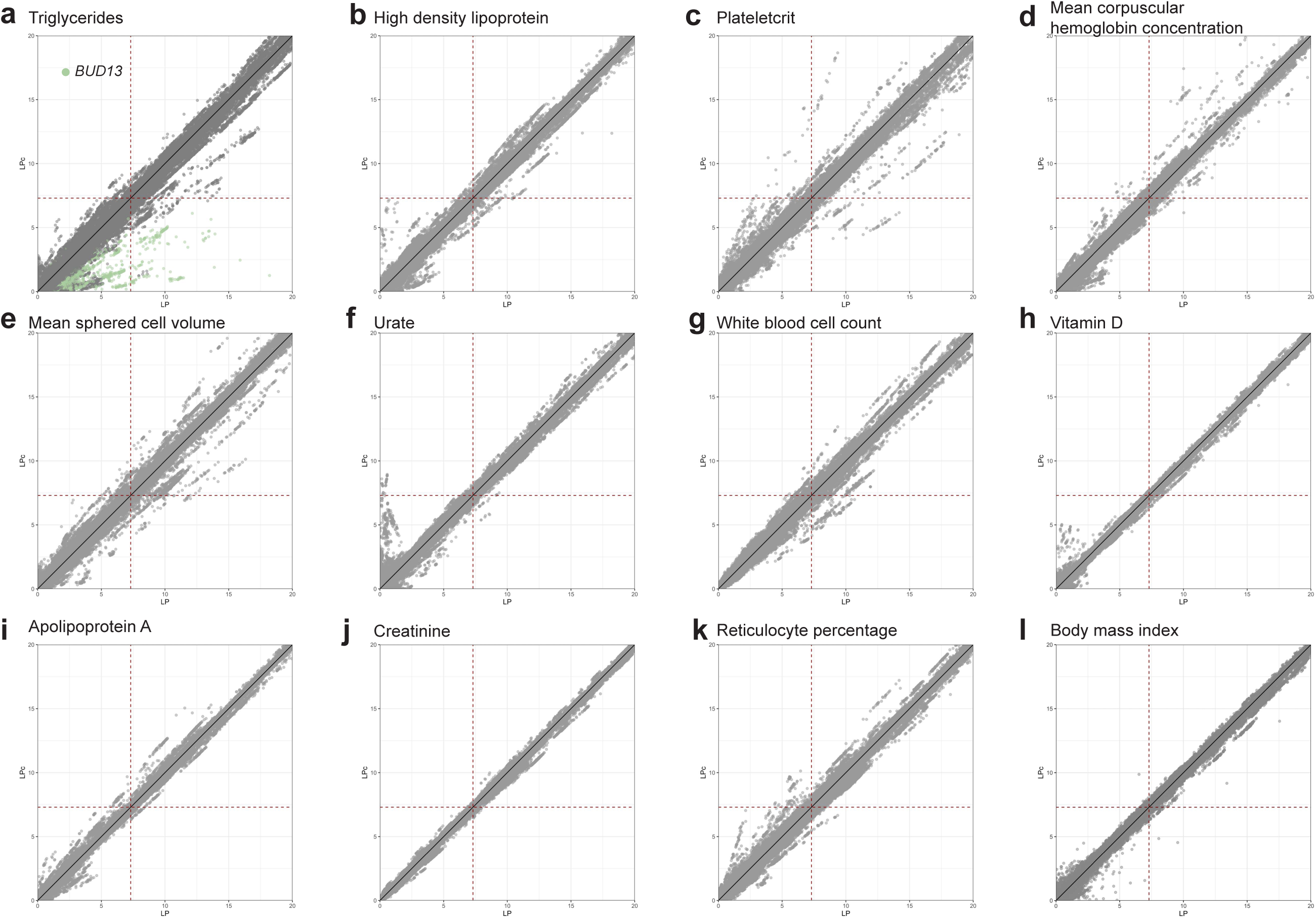
Multi-trait meta-analysis (mtCOJO) conditional analysis on additional significant forward Mendelian Randomization (MR) traits. Scatterplot of −log(*P*) from conditioning on significant MR traits using mtCOJO (LPc) vs. −log(*P*) from standard mLOY GWAS (LP) in MVP European ancestry. Red dashed lines indicate genome-wide significance at *P*<5×10^−8^. *BUD13* gene region is highlighted in trigylcerides (chr11:116,519,358-117,091,609).

**Supplementary Fig. 20.**
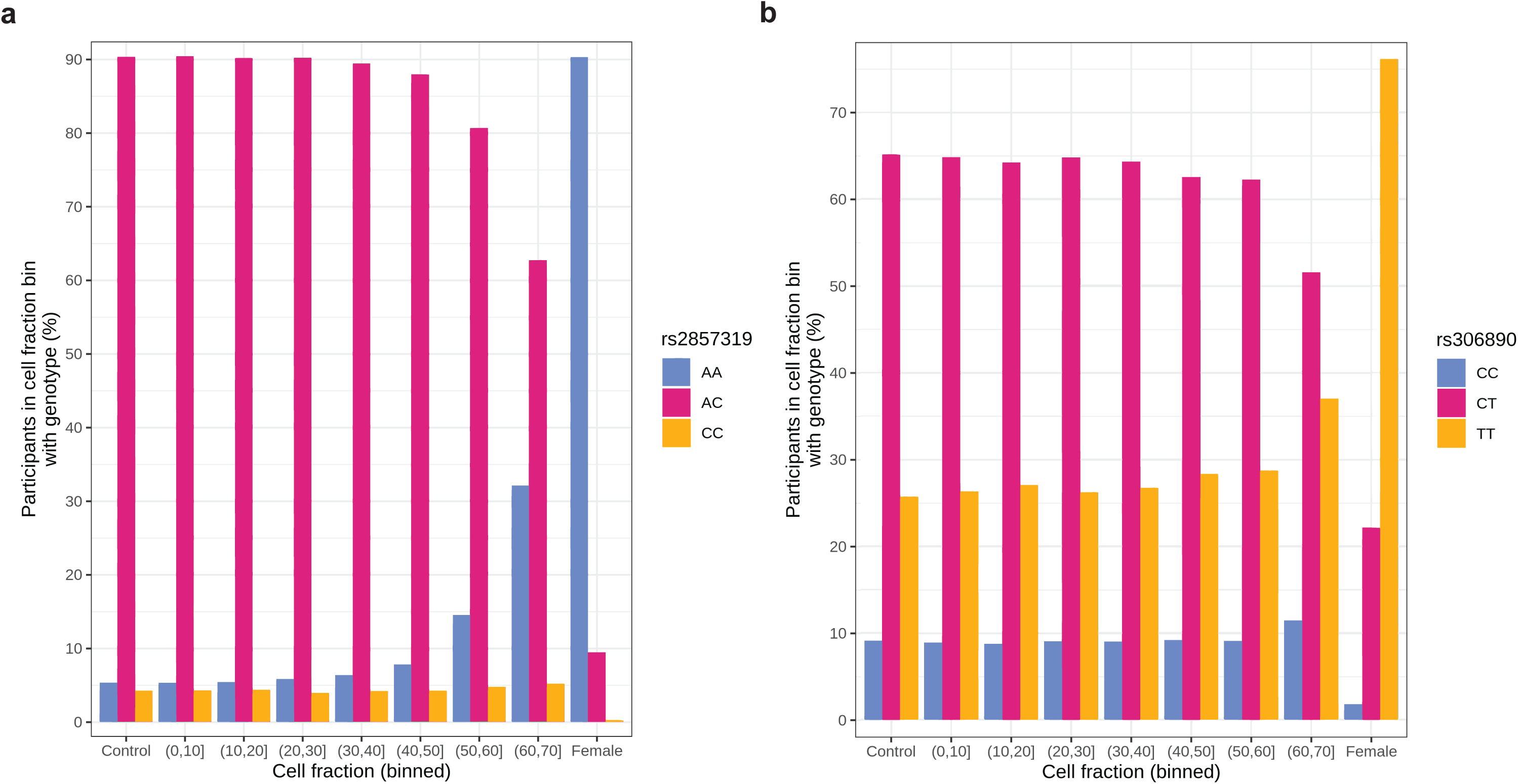
Rejected chromosome X associations. Two genome-wide significant variants on chromosome X, **(a)** rs2857319 and **(b)** rs306890, near the borders of PAR1 and PAR2, respectively, were found to be likely false positives. Plotting the percentage of participants with each genotype across cell fraction bins reveals a heterozygous dropout in higher cell fractions, suggesting genotyping error.

## Million Veteran Program: Consortium Acknowledgement for Manuscripts

### MVP Executive Committee

- Co-Chair: J. Michael Gaziano, M.D., M.P.H.
- Co-Chair: Rachel Ramoni, D.M.D., Sc.D.
- Jim Breeling, M.D. (ex-officio)
- Kyong-Mi Chang, M.D.
- Grant Huang, Ph.D.
- Sumitra Muralidhar, Ph.D.
- Christopher J. O’Donnell, M.D., M.P.H.
- Philip S. Tsao, Ph.D.

### MVP Program Office

- Sumitra Muralidhar, Ph.D.
- Jennifer Moser, Ph.D.

### MVP Recruitment/Enrollment

- Recruitment/Enrollment Director/Deputy Director, Boston
- Stacey B. Whitbourne, Ph.D.; Jessica V. Brewer, M.P.H.
- MVP Coordinating Centers

- Clinical Epidemiology Research Center (CERC), West Haven – John Concato, M.D., M.P.H.
- Cooperative Studies Program Clinical Research Pharmacy Coordinating Center, Albuquerque - Stuart Warren, J.D., Pharm D.; Dean P. Argyres, M.S.
- Genomics Coordinating Center, Palo Alto – Philip S. Tsao, Ph.D.
- Massachusetts Veterans Epidemiology Research Information Center (MAVERIC), Boston - J. Michael Gaziano, M.D., M.P.H.
- MVP Information Center, Canandaigua – Brady Stephens, M.S.
- Core Biorepository, Boston – Mary T. Brophy M.D., M.P.H.; Donald E. Humphries, Ph.D.
- MVP Informatics, Boston – Nhan Do, M.D.; Shahpoor Shayan
- Data Operations/Analytics, Boston – Xuan-Mai T. Nguyen, Ph.D.

### MVP Science

- Genomics - Christopher J. O’Donnell, M.D., M.P.H.; Saiju Pyarajan Ph.D.; Philip S. Tsao, Ph.D.
- Phenomics - Kelly Cho, M.P.H, Ph.D.
- Data and Computational Sciences – Saiju Pyarajan, Ph.D.
- Statistical Genetics – Elizabeth Hauser, Ph.D.; Yan Sun, Ph.D.; Hongyu Zhao, Ph.D.

### MVP Local Site Investigators

- Atlanta VA Medical Center (Peter Wilson) - Bay Pines VA Healthcare System (Rachel McArdle)
- Birmingham VA Medical Center (Louis Dellitalia)
- Cincinnati VA Medical Center (John Harley)
- Clement J. Zablocki VA Medical Center (Jeffrey Whittle)
- Durham VA Medical Center (Jean Beckham)
- Edith Nourse Rogers Memorial Veterans Hospital (John Wells)
- Edward Hines, Jr. VA Medical Center (Salvador Gutierrez)
- Fayetteville VA Medical Center (Gretchen Gibson)
- VA Health Care Upstate New York (Laurence Kaminsky)
- New Mexico VA Health Care System (Gerardo Villareal)
- VA Boston Healthcare System (Scott Kinlay)
- VA Western New York Healthcare System (Junzhe Xu)
- Ralph H. Johnson VA Medical Center (Mark Hamner)
- Wm. Jennings Bryan Dorn VA Medical Center (Kathlyn Sue Haddock)
- VA North Texas Health Care System (Sujata Bhushan)
- Hampton VA Medical Center (Pran Iruvanti)
- Hunter Holmes McGuire VA Medical Center (Michael Godschalk)
- Iowa City VA Health Care System (Zuhair Ballas)
- Jack C. Montgomery VA Medical Center (Malcolm Buford)
- James A. Haley Veterans’ Hospital (Stephen Mastorides)
- Louisville VA Medical Center (Jon Klein)
- Manchester VA Medical Center (Nora Ratcliffe)
- Miami VA Health Care System (Hermes Florez)
- Michael E. DeBakey VA Medical Center (Alan Swann)
- Minneapolis VA Health Care System (Maureen Murdoch)
- N. FL/S. GA Veterans Health System (Peruvemba Sriram)
- Northport VA Medical Center (Shing Shing Yeh)
- Overton Brooks VA Medical Center (Ronald Washburn)
- Philadelphia VA Medical Center (Darshana Jhala)
- Phoenix VA Health Care System (Samuel Aguayo)
- Portland VA Medical Center (David Cohen)
- Providence VA Medical Center (Satish Sharma)
- Richard Roudebush VA Medical Center (John Callaghan)
- Salem VA Medical Center (Kris Ann Oursler)
- San Francisco VA Health Care System (Mary Whooley)
- South Texas Veterans Health Care System (Sunil Ahuja)
- Southeast Louisiana Veterans Health Care System (Amparo Gutierrez)
- Southern Arizona VA Health Care System (Ronald Schifman)
- Sioux Falls VA Health Care System (Jennifer Greco)
- St. Louis VA Health Care System (Michael Rauchman)
- Syracuse VA Medical Center (Richard Servatius)
- VA Eastern Kansas Health Care System (Mary Oehlert)
- VA Greater Los Angeles Health Care System (Agnes Wallbom)
- VA Loma Linda Healthcare System (Ronald Fernando)
- VA Long Beach Healthcare System (Timothy Morgan)
- VA Maine Healthcare System (Todd Stapley)
- VA New York Harbor Healthcare System (Scott Sherman)
- VA Pacific Islands Health Care System (Gwenevere Anderson)
- VA Palo Alto Health Care System (Philip Tsao)
- VA Pittsburgh Health Care System (Elif Sonel)
- VA Puget Sound Health Care System (Edward Boyko)
- VA Salt Lake City Health Care System (Laurence Meyer)
- VA San Diego Healthcare System (Samir Gupta)
- VA Southern Nevada Healthcare System (Joseph Fayad)
- VA Tennessee Valley Healthcare System (Adriana Hung)
- Washington DC VA Medical Center (Jack Lichy)
- W.G. (Bill) Hefner VA Medical Center (Robin Hurley)
- White River Junction VA Medical Center (Brooks Robey)
- William S. Middleton Memorial Veterans Hospital (Robert Striker)

